# Perceptions of and obstacles to SARS-CoV-2 vaccination among adults in Lebanon: a cross-sectional online survey

**DOI:** 10.1101/2021.05.21.21257613

**Authors:** Nadeem E. Abou-Arraj, Diana Maddah, Vanessa Buhamdan, Roua Abbas, Nadine Jawad, Fatima Karaki, Nael H. Alami, Pascal Geldsetzer

## Abstract

The COVID-19 pandemic is an additional burden on Lebanon’s stressed population, fragmented healthcare system, and political, economic, and refugee crises. Understanding the population’s intentions to vaccinate, and perceptions of and obstacles to SARS-CoV-2 vaccination, can inform Lebanon’s vaccination efforts. We performed a cross-sectional study from 29 Jan 2021 to 11 Mar 2021 using an online questionnaire in Arabic via convenience “snowball” sampling to assess the perceptions of adults residing in Lebanon. 1,185 adults participated in the survey. % [95% CI: 43.2%-49.0%] of survey participants intended to take the SARS-CoV-2 vaccine when available to them, 19.0% [16.8%-21.4%] indicated that they would not, and 34.0% [31.3%-36.8%] were unsure. The most common reasons for hesitancy were concerns about safety, limited testing, side effects, and efficacy. Vaccine hesitancy appears to be high in Lebanon. Disseminating clear, consistent, evidence-based safety and efficacy information on vaccines may help reduce vaccine hesitancy, especially among the large proportion of adults who appear to be unsure about (rather than opposed to) vaccination.

## INTRODUCTION

As of 18 May 2021, Lebanon’s cumulative positive COVID-19 case count was 536,554 people (78,611 cases per million), and at least 7,641 people have died [1]. Already a devastating toll, this number is likely significantly underreported due to lack of testing and fragmented health infrastructure in the country [2]. In addition to the pandemic, Lebanon has been struggling with multiple challenges: a political crisis and economic collapse driven by corruption that began in 2019 and worsened throughout 2020, leading to widespread mistrust of the government, inflation, unemployment, poverty, and increased food insecurity, on top of the strain of being the country with the highest number of refugees per capita in the world due to the protracted Syrian refugee crisis [2–6]. Given Lebanon’s compounding crises and limited resources, mass vaccination is a challenging but vital mission. Understanding the population’s perceptions of SARS-CoV-2 vaccines is critical for implementing a successful vaccination campaign in the country, providing extra support for vulnerable populations, and bolstering demand for vaccination.

SARS-CoV-2 vaccination began in Lebanon on 14 February 2021 [7]. Despite comprehensive planning and outreach efforts from the Lebanese Ministry of Public Health and prominent non-governmental organizations including the World Health Organization and UNICEF, and international assistance from the World Bank and COVAX to secure adequate supply of SARS-CoV-2 vaccines of multiple formulations, as of 18 May 2021, only 208,086 people had been fully vaccinated, representing approximately 3.1% of the total population [1,8– 10]. Inadequate demand, possibly due to vaccine hesitancy and/or logistical challenges, appears to be at least part of the reason for slow vaccination; at the largest public hospital in Beirut, up to 30% of vaccination appointments went unfilled during days in April 2021 [4]. As of 19 May 2021, only roughly 20% of the population has registered for the vaccine using the online national vaccine registration tool through the Ministry of Public Health [7]. Given the significant proportion of the population still needing vaccination, understanding perceptions of the vaccines and perceived obstacles is important to increasing vaccination rates to end the pandemic.

Previous studies of vaccination in Lebanon provide some background for anticipating the response to SARS-CoV-2 vaccination. An online survey of Lebanese in April-May 2020 found that 69% of the convenience sample (predominantly young, university-educated, and male) indicated they would take a hypothetical vaccine against SARS-CoV-2, though at that time, none had been developed or approved [11]. One study of seasonal influenza vaccination in Lebanon demonstrated that 28% of ambulatory adult patients at pharmacies were vaccinated (rates comparable to influenza vaccination across Europe) and factors associated with vaccination included belief in vaccine necessity, efficacy, and safety, as well as having private insurance, elderly age, higher educational attainment, and higher physical activity [12,13]. In a study of Lebanese physicians’ perspective on the pneumococcal and influenza vaccines, physicians cited availability and cost concerns, patients’ declining to be vaccinated, and physicians’ doubts over efficacy as barriers to vaccination [14]. For the refugee population specifically, several studies have assessed vaccination in displaced Syrians living in Lebanon, finding inadequate rates of childhood vaccination, high variability by location within Lebanon, and challenges in maintaining vaccination records [15–17].

Studies of SARS-CoV-2 vaccination intentions and perceptions in other countries and regions can also be helpful in informing the vaccination campaign in Lebanon. Surveys assessing SARS-CoV-2 vaccine hesitancy around the world cite reasons for hesitancy including safety concerns, worries about side effects, doubts about efficacy, unfavorable personal risk/benefits assessments of the SARS-CoV-2 vaccine, as well as general mistrust in science and government [18–20]. Other barriers include distribution and uptake of the vaccines at scale, which are matters of logistics, healthcare access, and public perception of the vaccine [21,22]. These issues may be exacerbated in the Lebanese context given the deeply seated mistrust of government as well as compounding social crises, but to our knowledge, no such study has been done to elucidate perceptions of the SARS-CoV-2 vaccination campaign in Lebanon.

It is therefore important to establish an evidence-based understanding of the perceptions of the SARS-CoV-2 vaccines among the Lebanese population. Our study’s aims were 1) to assess rates of intention to vaccinate and vaccine hesitancy in Lebanon; 2) to determine how vaccine hesitancy in Lebanon varies by sociodemographic, economic, and geographic characteristics; and 3) to understand individuals’ motivations for vaccinating and concerns and obstacles to vaccination.

## METHODS

### Study Design

To assess perceptions of SARS-CoV-2 vaccination in Lebanon, we designed a cross-sectional descriptive study employing a remote online Arabic survey. We originally intended to distribute it to randomly selected Lebanese phone numbers to obtain an unbiased nationally representative sample, but during piloting, this method needed to be aborted because of exceedingly low response rates (less than 1%), thought to be due to mistrust of messages and links received from an unknown phone number. Given these constraints, we changed our distribution methods to convenience “snowball” sampling, a method further described below that had successfully been used elsewhere in the Middle East to quickly recruit a large sample size [23].

After making these adjustments, an anonymous, online, self-administered survey was created and distributed using convenience “snowball” sampling. The recruitment and survey period was six weeks: 29 January 2021 - 11 March 2021. This time period spanned before and after initiation of SARS-CoV-2 vaccination in Lebanon, which began on 14 February 2021 [7].

### Study Population

The target population was all adults living in Lebanon, including the significant refugee population. While all adults living in Lebanon were eligible to participate, they needed to be able to access the self-administered online survey tool, either on a mobile phone or computer. The survey was in Arabic, so participants were required to be literate in Arabic or to be assisted by someone who was.

### Recruitment and Sampling

We created an anonymous online Qualtrics survey (see appendix) distributed and administered in Arabic [24]. To disseminate the survey, we created a recruitment message in Arabic (see appendix) that introduced the survey and asked if the message recipients would be interested in participating. Given the convenience “snowball” sampling method, the research team initiated recruitment by sending the survey to their contacts and throughout their organizations. Participants who were interested in the study proceeded to the survey via a link at the end of the recruitment message. The recruitment message also invited participants to forward the message to their contacts. The message could be sent through WhatsApp, SMS, social media, and email. There was no follow up to determine whether individuals who received the recruitment message completed or forwarded the survey. No incentives were provided to participants for participation in the study.

### Survey Tool and Data Collection

The survey was created using the Qualtrics online survey platform [24]. It was anonymous, self-administered, and did not require more than basic literacy in Arabic.

The survey content was created by the research team (the majority of whom were fluent in Arabic and English), using several other SARS-CoV-2 vaccine perception studies as a guide [25,26], and tailored to the Lebanese context. It was piloted in small groups of contacts of the research team who had varying educational backgrounds and health literacy and revised several times prior to official survey launch.

The survey consisted of 31 multiple-choice and free-response questions (depending on branch points, participants were not asked each question), divided into an introduction with the informed consent document, followed by questions about screening, demographics, questions experience with COVID-19, and perceptions of SARS-CoV-2 vaccination. No identifying data was collected. The first questions asked participants to provide informed consent, to affirm that they were 18 years or older and living in Lebanon, and to verify that they had heard of “coronavirus” (as SARS-CoV-2 and COVID-19 are referred to in Lebanese Arabic). If they declined, the survey automatically ended. If they passed these questions, they were able to proceed with the survey.

The survey consisted of three sections. The “coronavirus vaccine” section assessed intentions to receive or not to receive vaccination and motivations behind these decisions. It also asked about participants’ preferred location to receive the coronavirus vaccine, how much money they would be willing to pay for the vaccine if it were to cost money, and if monetary incentives could influence their decision on taking the vaccine.

The “experience with coronavirus” section surveyed participants’ most frequently used news sources about coronavirus, their most trusted news sources, how often they wore a face mask as a preventative measure when leaving their homes, and whether they or anyone they knew had been infected with coronavirus.

The “demographics” section included questions on gender, age, religion, household income for the year 2019, education level, geographic location, nationality, and refugee status. Once participants started the survey, they had 48 hours to complete it before the survey automatically recorded their responses. Participants were prevented from taking the survey multiple times on the same device using a Qualtrics feature based on browser cookies.

### Data Analysis

Data were downloaded from Qualtrics as a .csv file and translated into English using Microsoft Excel [27]. Data cleaning and statistical analysis were performed in R computer software and focused on description rather than identifying causal links [28].

Several quality measures were implemented. To ensure that participants had at least a baseline familiarity with the survey topics, one of the screening questions asked whether participants had heard of “coronavirus.” If they had not, they were not able to proceed with the survey; only 5 participants (0.42%) were excluded because of not being aware of “coronavirus.” To identify participants who randomly clicked through the survey, a filter was applied to detect participants who completed the survey in less than 120 seconds; no participants did so.

#### Quantitative Data Analysis

For some multiple-choice questions, similar categorical responses were consolidated into binary or fewer categories to facilitate interpretation. For binary and categorical variables, the absolute number and relative proportions of participants who selected each response was calculated. Wilson score 95% confidence intervals were calculated for proportions.

Given that the survey period spanned the initiation of SARS-CoV-2 vaccination in Lebanon, a sub-analysis was performed in which participants were divided by whether they completed the survey before or after vaccine initiation. For each of these sub-groups, sample demographics were recalculated and are displayed in Table 1. We did not use sampling weights in our analysis given that this was a non-probabilistic sample of the Lebanese population that was unlikely to be representative of the general population even after weighting.

**Table 1:**
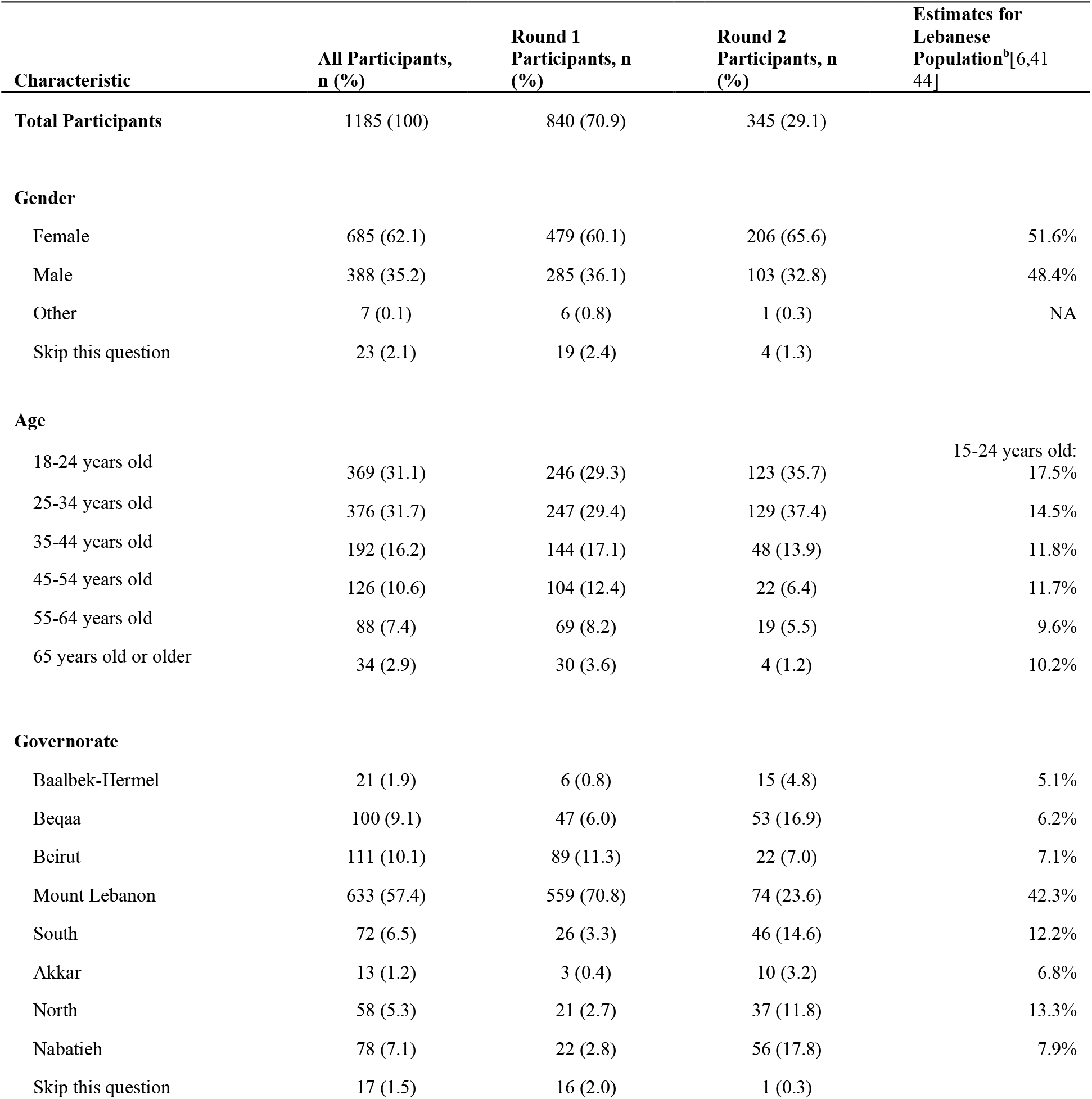
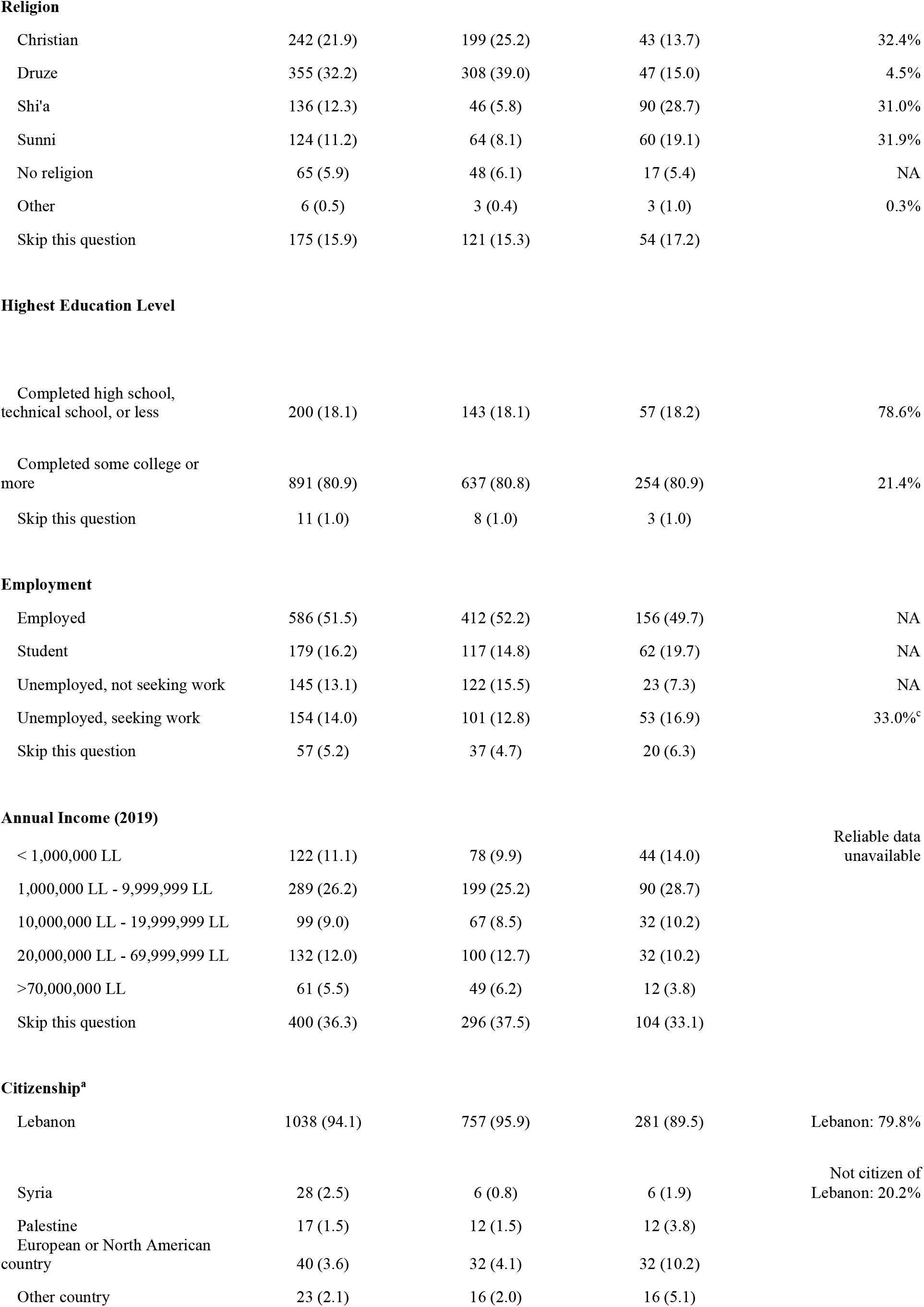
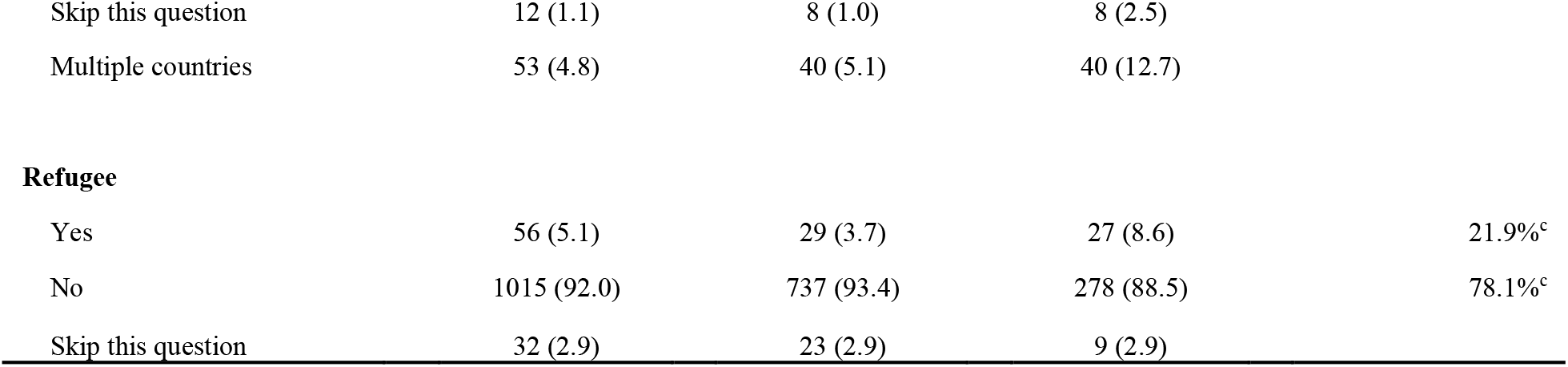
Participant Characteristics. This includes all who passed screening. The first column includes all participants aggregated. The second column describes those who completed the survey in the first round, prior to initiation of vaccination in Lebanon on 13 February 2021. The third column describes those who completed the survey in the second round, after initiation of vaccination in Lebanon. Because participants were not forced to answer all questions, this results in a different denominator for each question. LL: Lebanese Lira NA: Not available in data source ^a^Participants could select multiple answers. Proportions were calculated using a denominator of all participants who selected an answer for the question. ^b^Unless otherwise noted, estimates were obtained from government source that excluded refugees and used 4.84 million (2018) as total population. ^c^Estimates were obtained from source that included refugees and used 6.86 million (2020) as total population.

#### Qualitative Data Analysis

If participants indicated that they intend to vaccinate, they were then asked, “Why do you plan to get the coronavirus vaccine if and when it becomes available to you?” If they intended not to vaccinate, they were asked, “Why do you plan on not getting the coronavirus vaccine if and when it becomes available to you?” If they were uncertain about vaccinating, they were asked, “Why are you unsure about getting the coronavirus vaccine if and when it becomes available?”

These open-ended responses were coded using a uniform protocol in order to facilitate analysis (see appendix). First, each response was translated from Arabic into English by one of six bilingual members of the research team. This translation was then verified by a second bilingual member of the research team. The English translations were then coded using thematic analysis by one of eight members of the research team. A second coder reviewed the first team member’s codes to verify accuracy and authenticity. Disagreements were discussed and consensus was achieved usually between the two coders. Rarely, a third member of the coding team (NEA or VB) was needed to resolve a disagreement. The list of codes was constructed and updated iteratively throughout the coding process. Multiple codes could apply to the same response. To ensure consistency in coding between coders and responses, one team member (NEA) reviewed all codes. Codes were then ordered from most common to least common.

## RESULTS

### Sample Characteristics

1,390 participants initiated the survey. Of these, 1,185 (85.3%) provided informed consent and passed screening to begin the main survey (Figure 1). Among this group, 1,103 (93.1%) participants completed the entire survey. Sample demographic characteristics of those who passed screening are summarized in Table 1.

**Figure 1:**
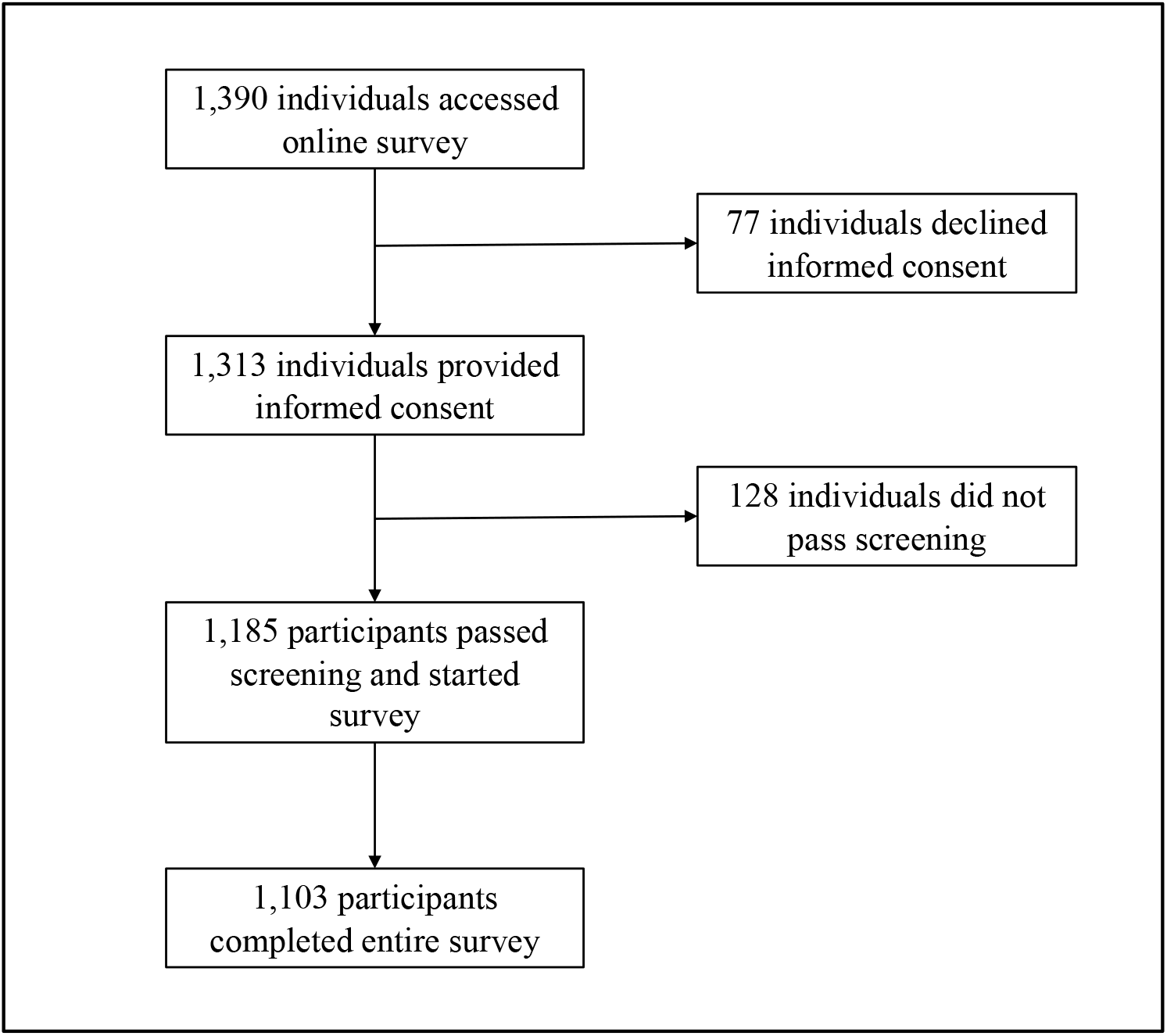
Survey Recruitment and Outcomes.

Compared to demographics of residents of Lebanon in general, our sample population had higher representation of individuals who were young, female, well-educated, identified with the Druze religion, and from the Mt. Lebanon region (Table 1). Underrepresented in our sample were the elderly, refugees, non-Lebanese citizens, members of the Sunni and Shi’a religions, and individuals from the less populated governorates of Akkar, Baalbek-Hermel, Nabatieh, North, and South.

### Quantitative Analysis

#### Intentions to Vaccinate

We found that 46.1% [95% CI: 43.2%-49.0%] of our survey participants intended to take the SARS-CoV-2 vaccine when available to them. 19.0% [16.8%-21.4%] indicated that they would not get vaccinated, and 34.0% [31.3%-36.8%] were unsure about vaccination (Table 2).

**Table 2:**
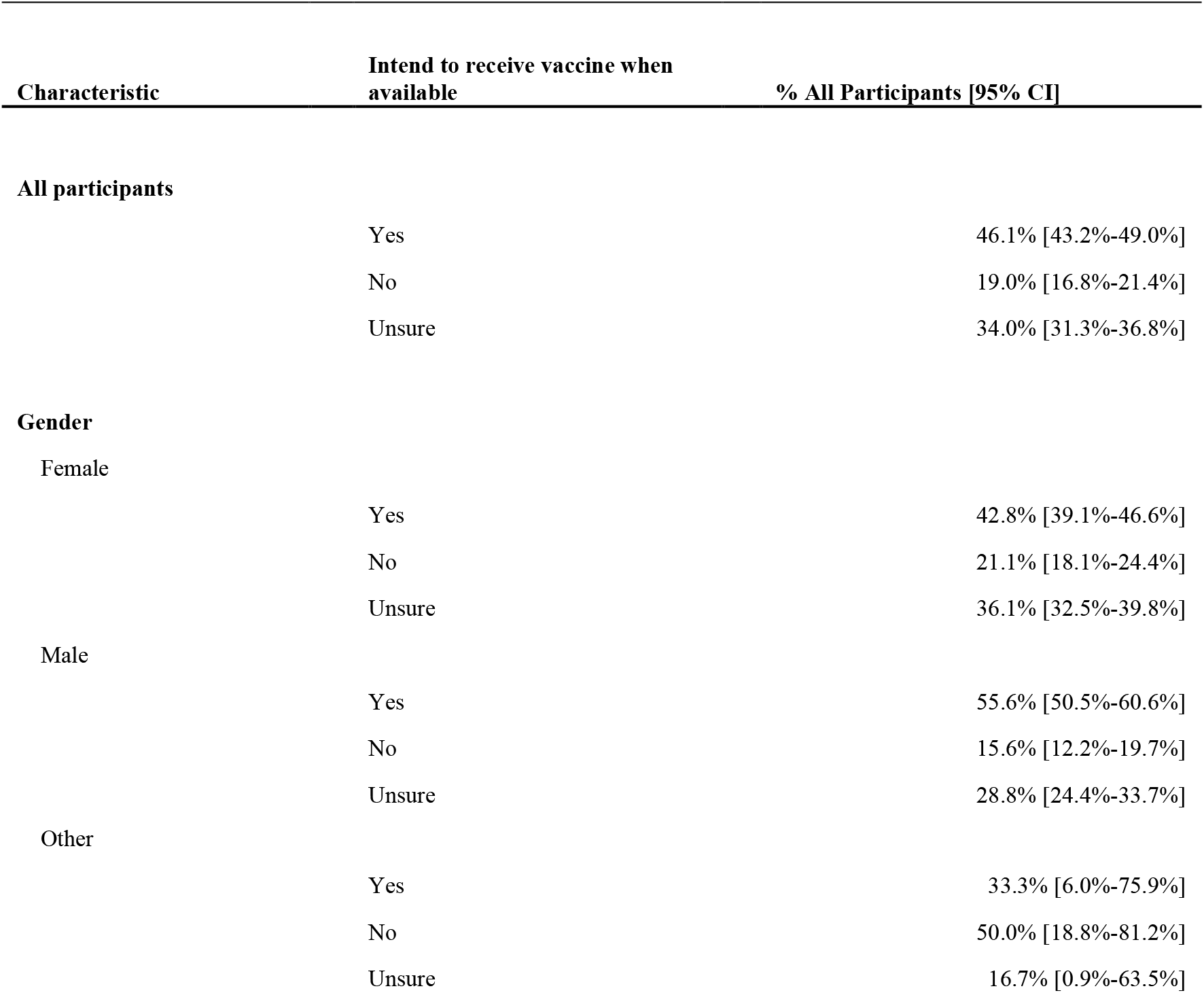
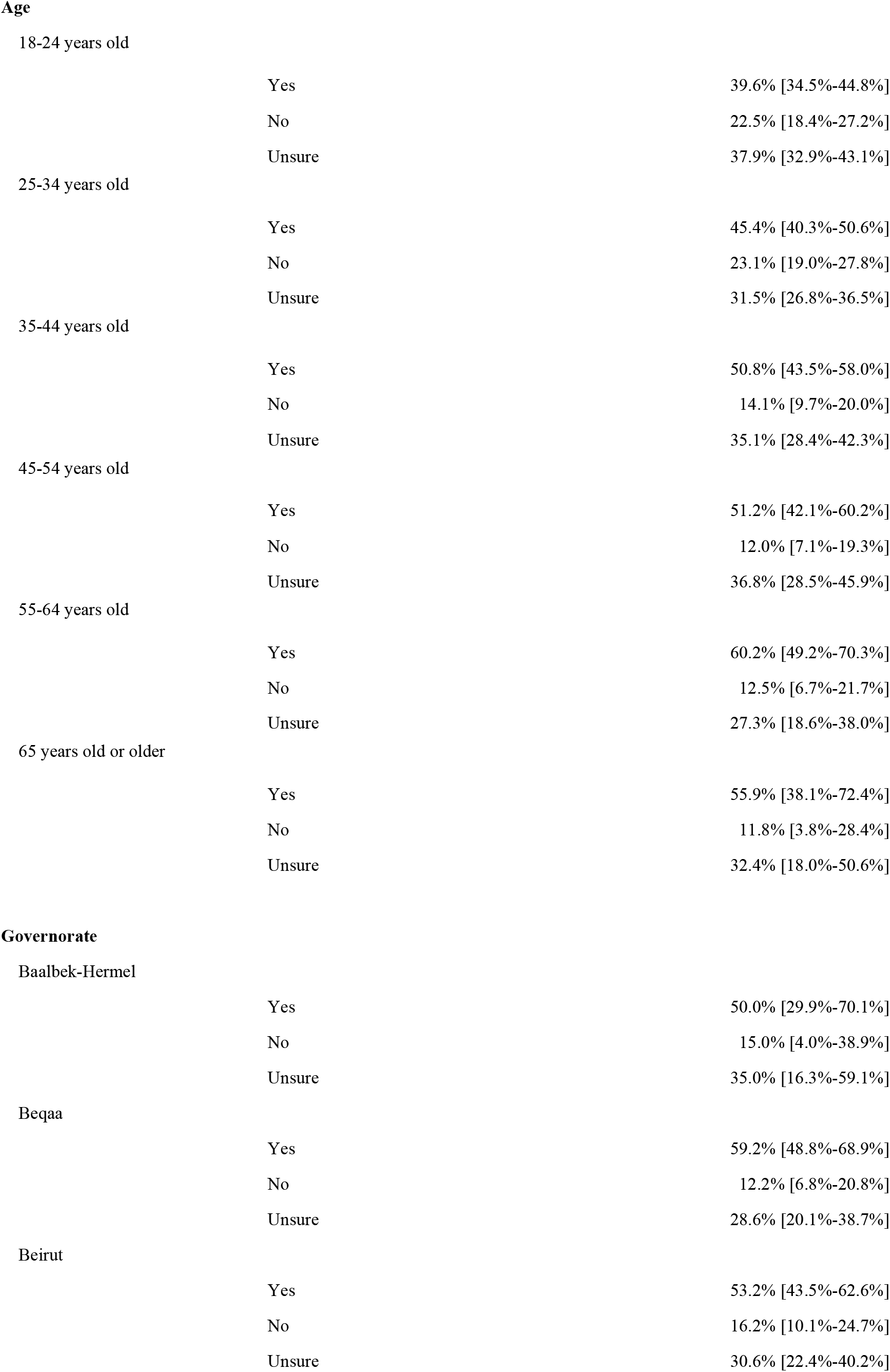
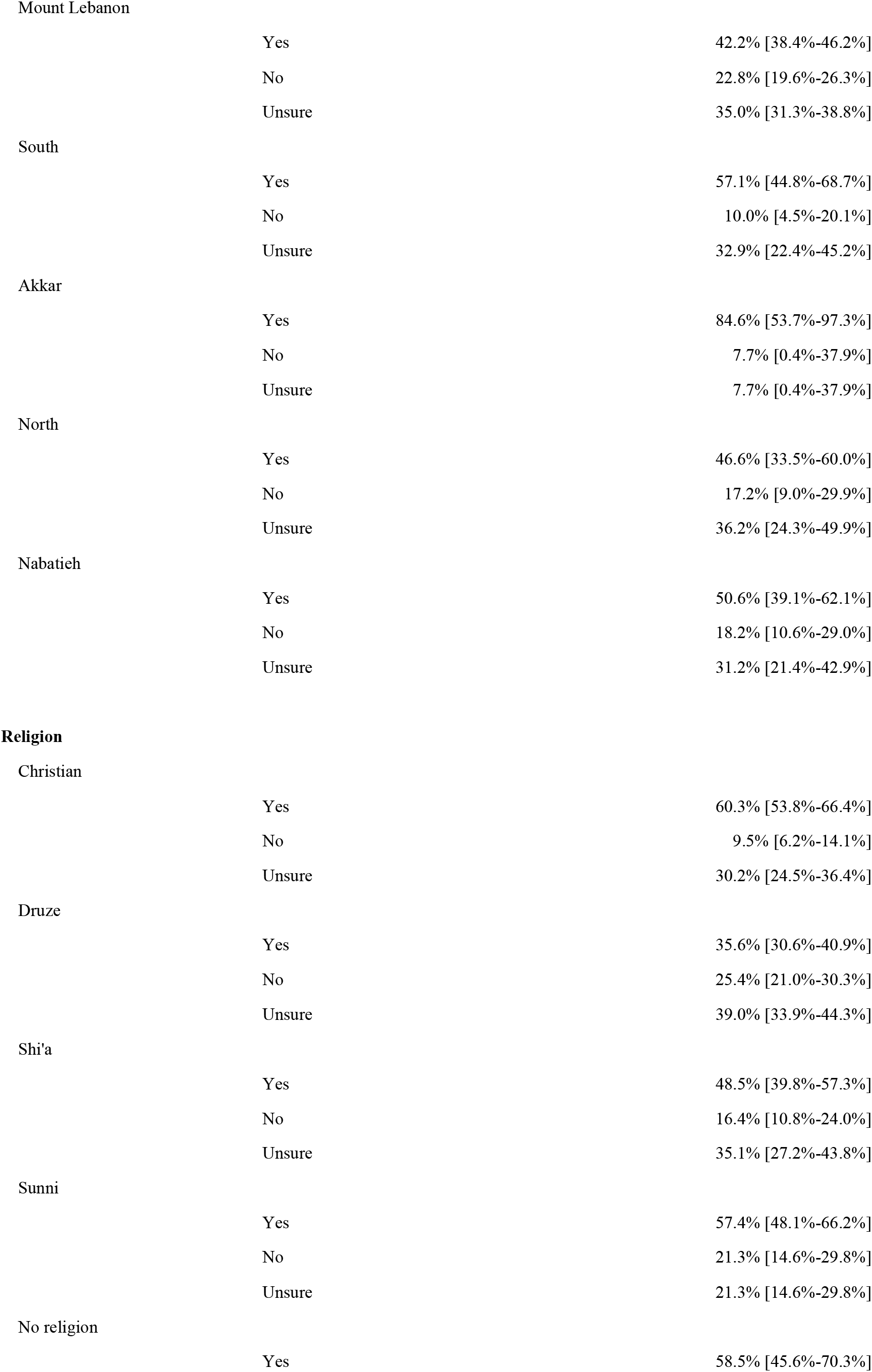
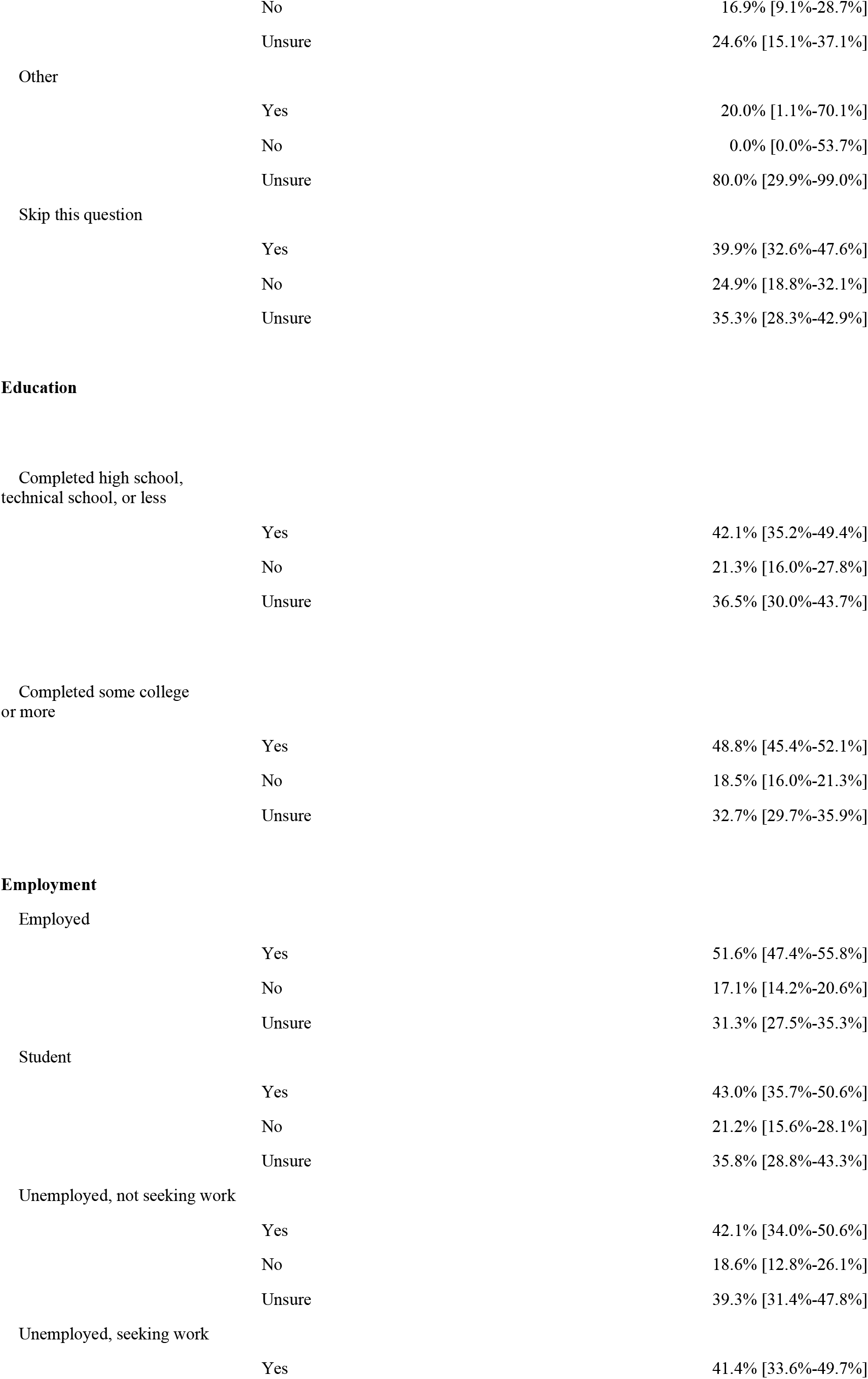
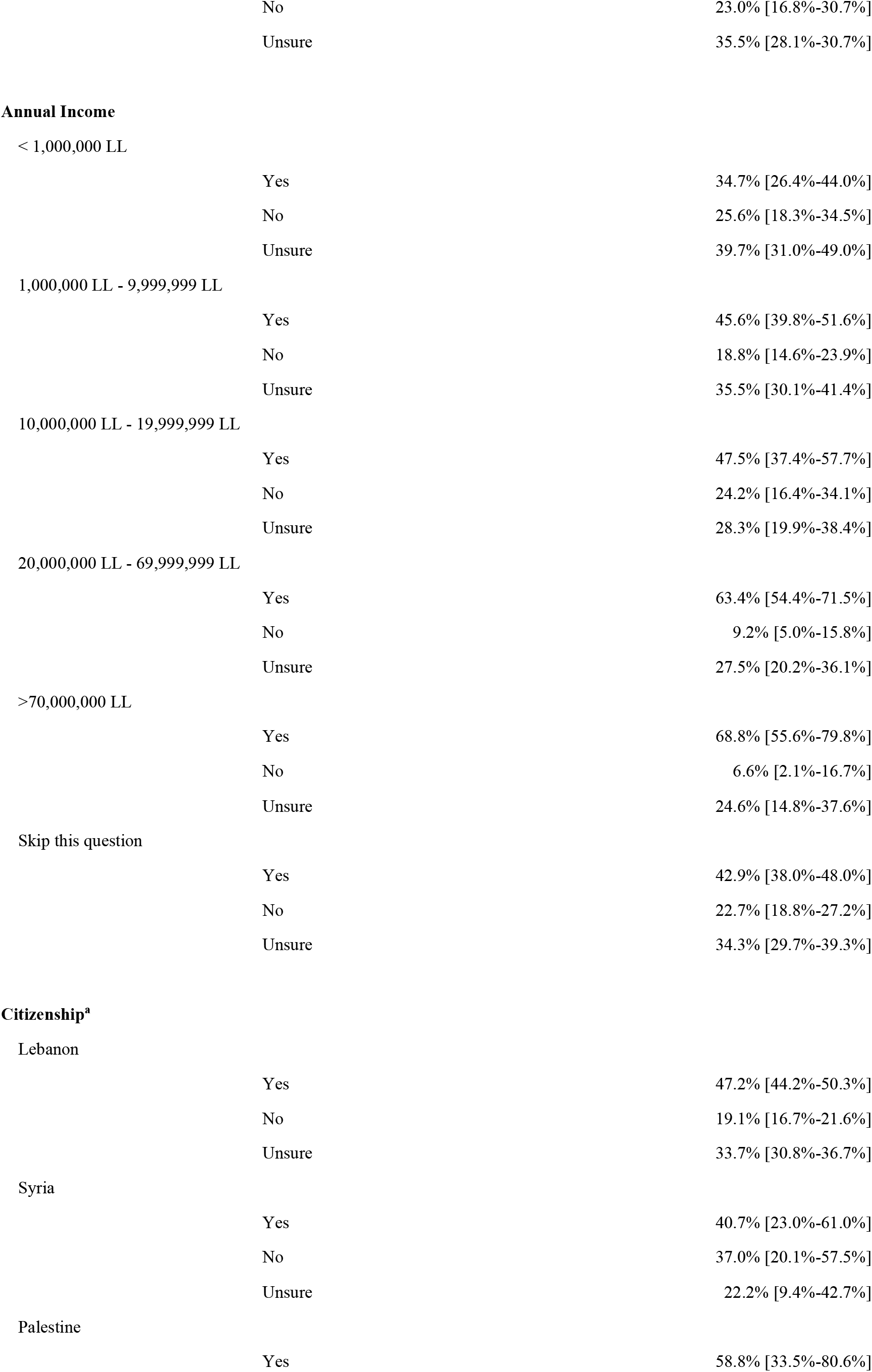
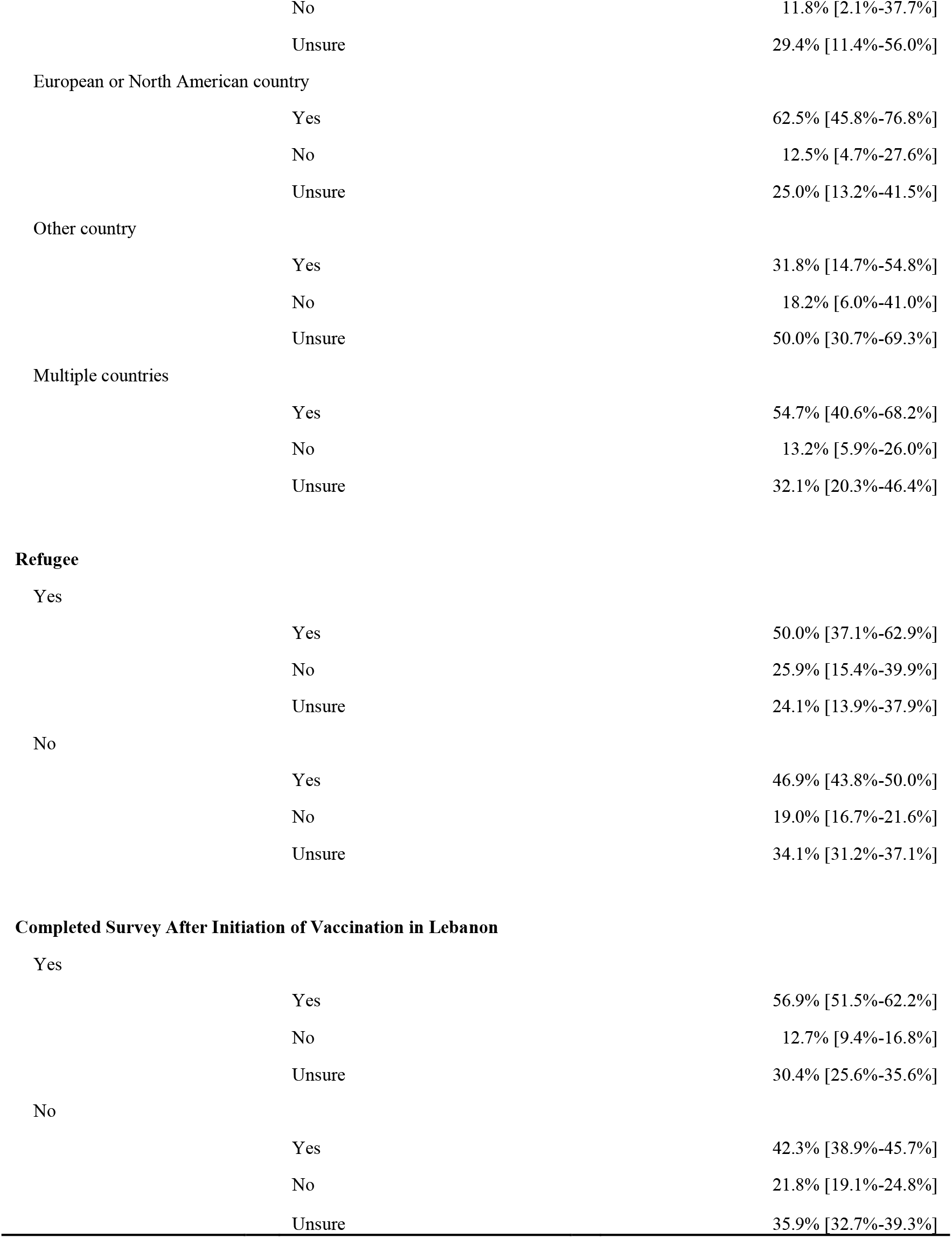
Intentions about vaccination by sociodemographic characteristics. This table shows intention to vaccinate by demographic characteristic for everyone who answered the intention to vaccinate question. For the analysis of each characteristic, we omitted participants who skipped the question, unless >10% of participants for that question skipped the question, in which case those who skipped the characteristic question were included in the analysis. We then calculated the proportion of each characteristic subcategory by intention to vaccinate, calculating Wilson Score confidence intervals. ^a^The survey allowed participants to choose multiple answers for this characteristic; consequently, the sum of all subcategories does not equal the number of all participants who answered the question.

#### Intentions to Vaccinate by Demographic Characteristics

Overall, participants were more likely to intend to vaccinate if they identified as male; lived in the Beqaa governorate (Mount Lebanon as reference); were a member of Christian, Sunni, or no religion (Druze as reference); and had higher household income (Table 2 & Figure 2).

**Figure 2:**
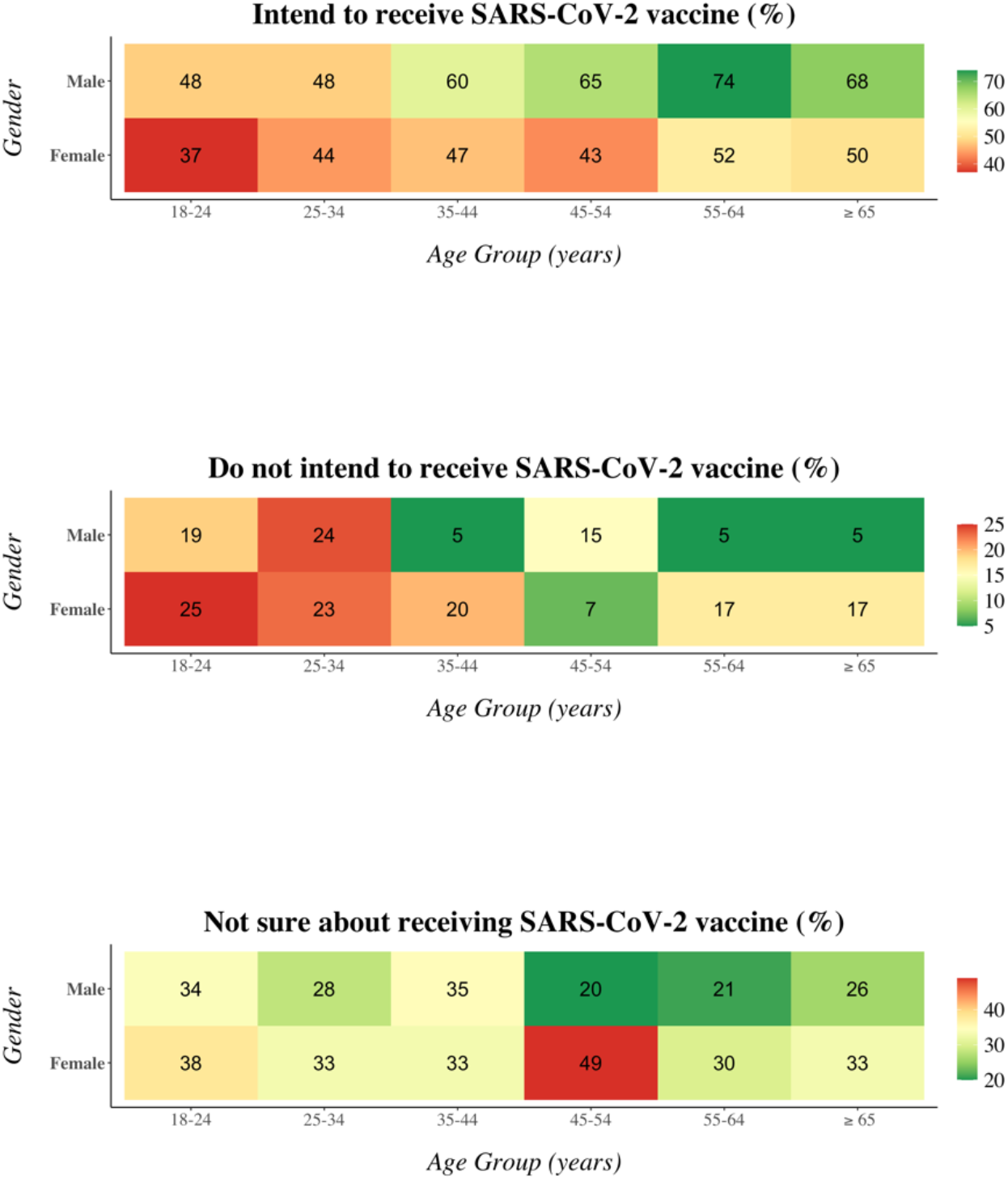
Intention to receive SARS-CoV-2 vaccine by Age Group and Gender. In these graphs, the sample population is stratified by age group and gender. For each stratum, the proportion (as a percentage) that indicated that they intend to receive the vaccine, do not intend to receive the vaccine, and are unsure about receiving the vaccine is displayed in the corresponding cell in the top, middle, and bottom graphs, respectively. This analysis only includes participants who provided age, gender, and their intentions about vaccination. As only 7 participants identified as “other” gender, only participants identifying as “male” or “female” were included.

There were less strong but still apparent trends toward higher proportions of participants’ intending to vaccinate if they identified as older in age; lived in Beirut or Akkar governorates (Mount Lebanon as reference); had attained higher educational status; or were employed. There were no apparent differences or trends in intention to vaccinate by country of citizenship or whether participants identified as refugees.

#### Intentions to Vaccinate by Experiences with COVID-19

Comparing participants who reported a personal history of COVID-19 infection (n = 321 [28.9%]) to those who did not (n = 790 [71.1%]), there did not appear to be large differences in vaccine acceptance (48.8% [45.2%-52.3%] vs. 42.9% [37.5%-48.6%]) (Table 3). However, those with a family member or close friend who had contracted COVID-19 (n = 1056 [94.2%]) were substantially more likely to intend to vaccinate, compared to individuals who did not (48.0% [44.9%-51.0%] vs. 33.3% [22.2%-46.4%]). Notably, the proportion of participants who did not have a close acquaintance who had been infected was low (n = 65 [5.8%]).

**Table 3:**
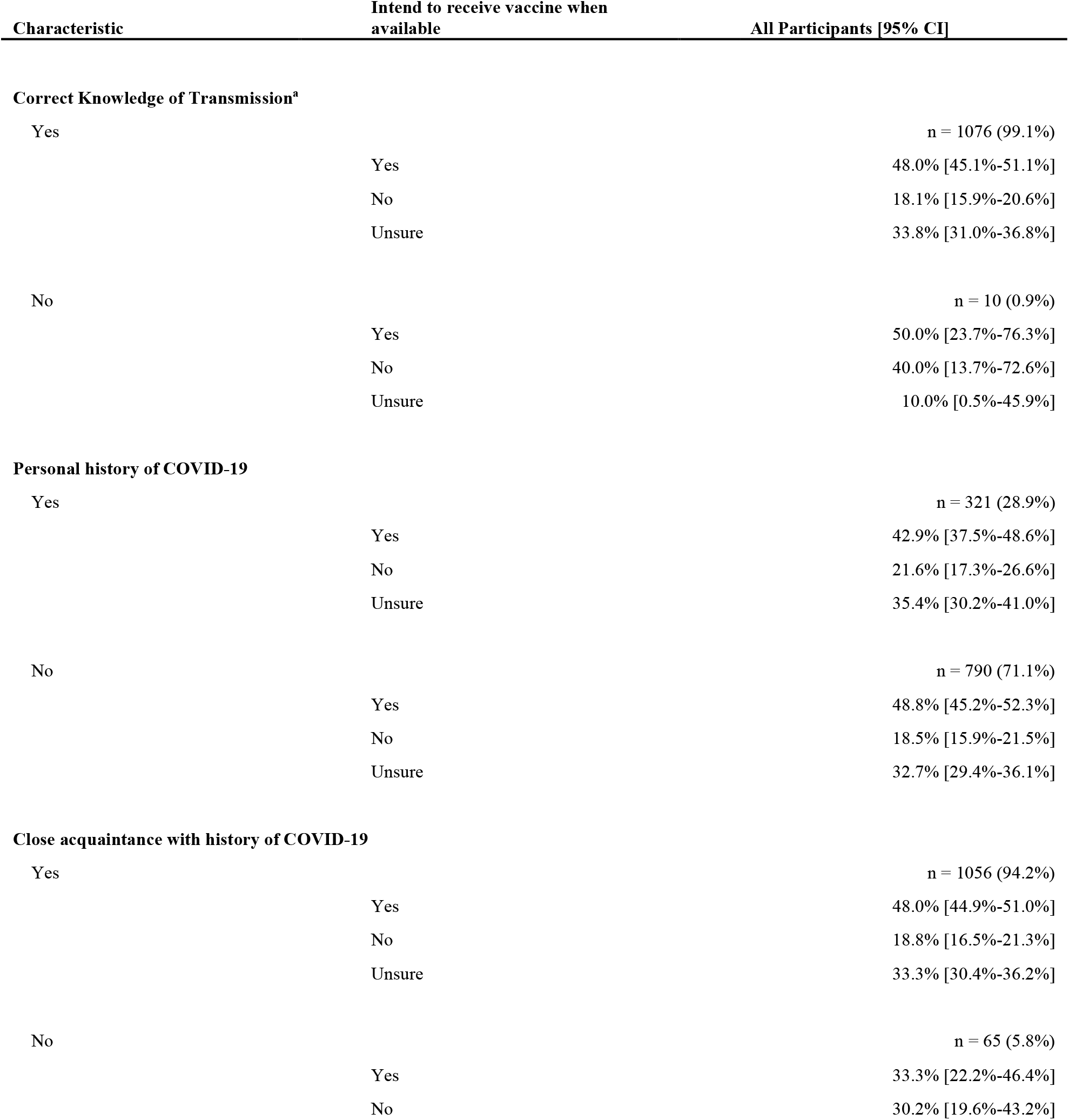
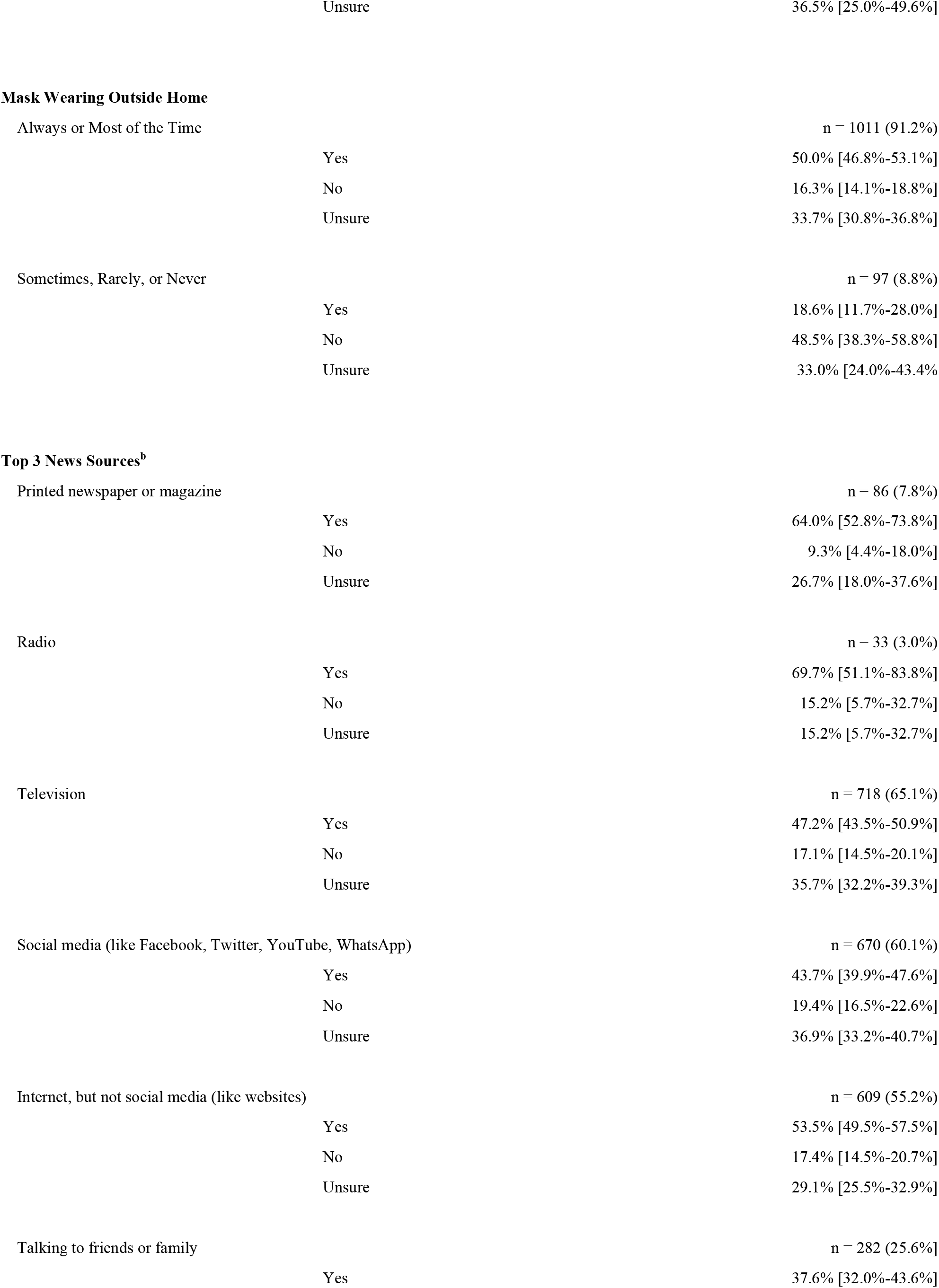
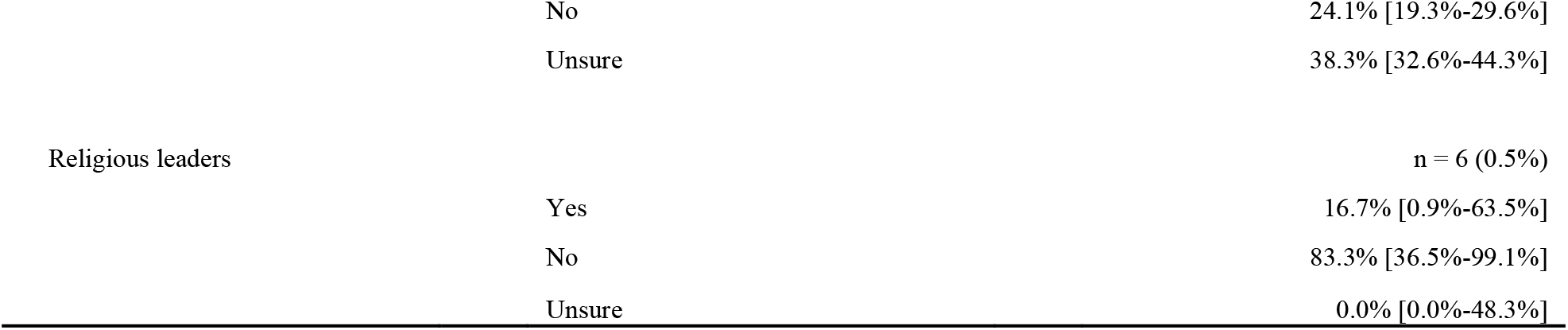
Intentions about vaccination by experiences with COVID-19. This shows intentions to vaccinate broken down by knowledge of and experience with COVID-19 for everyone who answered the intention to vaccinate question. For the analysis of each characteristic, we omitted participants who skipped the question, unless >10% of participants for that question skipped the question, in which case those who skipped the characteristic question were included in the analysis. We then calculated the proportion of each characteristic subcategory by intention to vaccinate, calculating Wilson Score confidence intervals. ^a^”Correct Knowledge” indicated a correct response to a multiple choice question asking, “In your understanding, how does someone become infected with coronavirus? Choose the best single answer.” The correct response was “Being physically close to an infected person.” Incorrect responses were “Eating raw food or untreated water,” and “Being bitten by an insect.” ^b^The survey allowed participants to choose multiple answers for this characteristic; consequently, the sum of all subcategories does not equal the number of all participants who answered the question.

When compared to participants who reported “sometimes,” “rarely,” or “never” wearing a mask when outside the home (n = 97 [8.8%]), participants who reported wearing a mask “always” or “most of the time” (n = 1011 [91.2%]) were much more likely to intend to vaccinate (50.0% [46.8%-53.1%] vs 18.6% [11.7%-28.0%]).

We also asked participants to identify their top three most commonly used sources of news for coronavirus. Participants who reported commonly obtaining news from newspapers and magazines or the radio were more likely to intend to vaccinate than those who reported television, social media, other internet websites, and family and friends as common sources of news. Very few participants (n = 6 [0.5%]) reported that religious leaders are a commonly used source of news.

#### Intentions to Vaccinate before and after Initiation of Vaccination in Lebanon

Those who completed the survey after initiation of vaccination were more likely to intend to vaccinate (56.9% [51.5%-62.2%]) when compared to those who completed the survey before initiation of vaccination (42.3% [38.9%-45.7%]) (Table 2). Importantly, these groups were contacted differently and differed in several key demographic characteristics: the group of participants who responded after initiation of vaccination was younger and had a higher proportion of participants that lived outside of the Mount Lebanon region, identified as Shi’a or Sunni rather than Druze or Christian, had European or North American or multiple citizenships, and identified as refugees (Table 1).

#### Logistical Considerations about Vaccination

The most commonly selected preferred locations to get vaccinated were hospitals, doctors’ offices, primary health centers, and pharmacies (Figure 3). Temporary vaccination sites were not popular. The most common sources of news about coronavirus were television, social media, and other internet websites (Figure 4). Participants trusted television and internet websites more than social media. Among participants who did not intend to vaccinate or were uncertain about vaccination, less than 2% stated that a monetary incentive would persuade them to become vaccinated.

**Figure 3:**
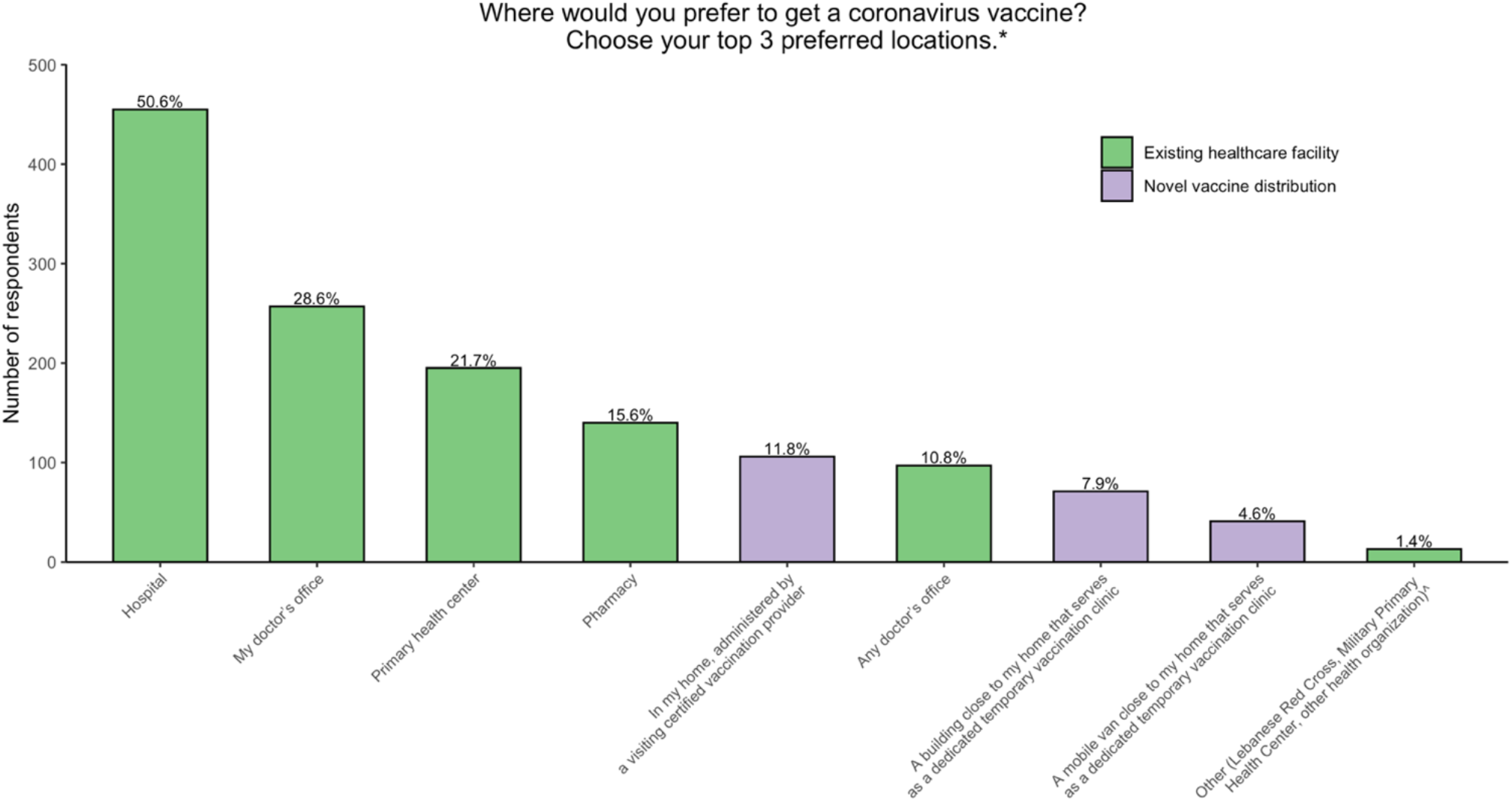
Preferred vaccination locations. Percentages of each response were calculated by dividing the number of participants who selected a response by the total number of participants who answered this question. ^*^The survey allowed participants to choose multiple answers for this question; consequently, the sum of all subcategories does not equal the number of all participants who answered the question. ^^^If participants selected “Other,” they could write in their own response. The three responses in parenthesis (Lebanese Red Cross, Military Primary Health Center, other health organization) encompass the written-in responses.

**Figure 4:**
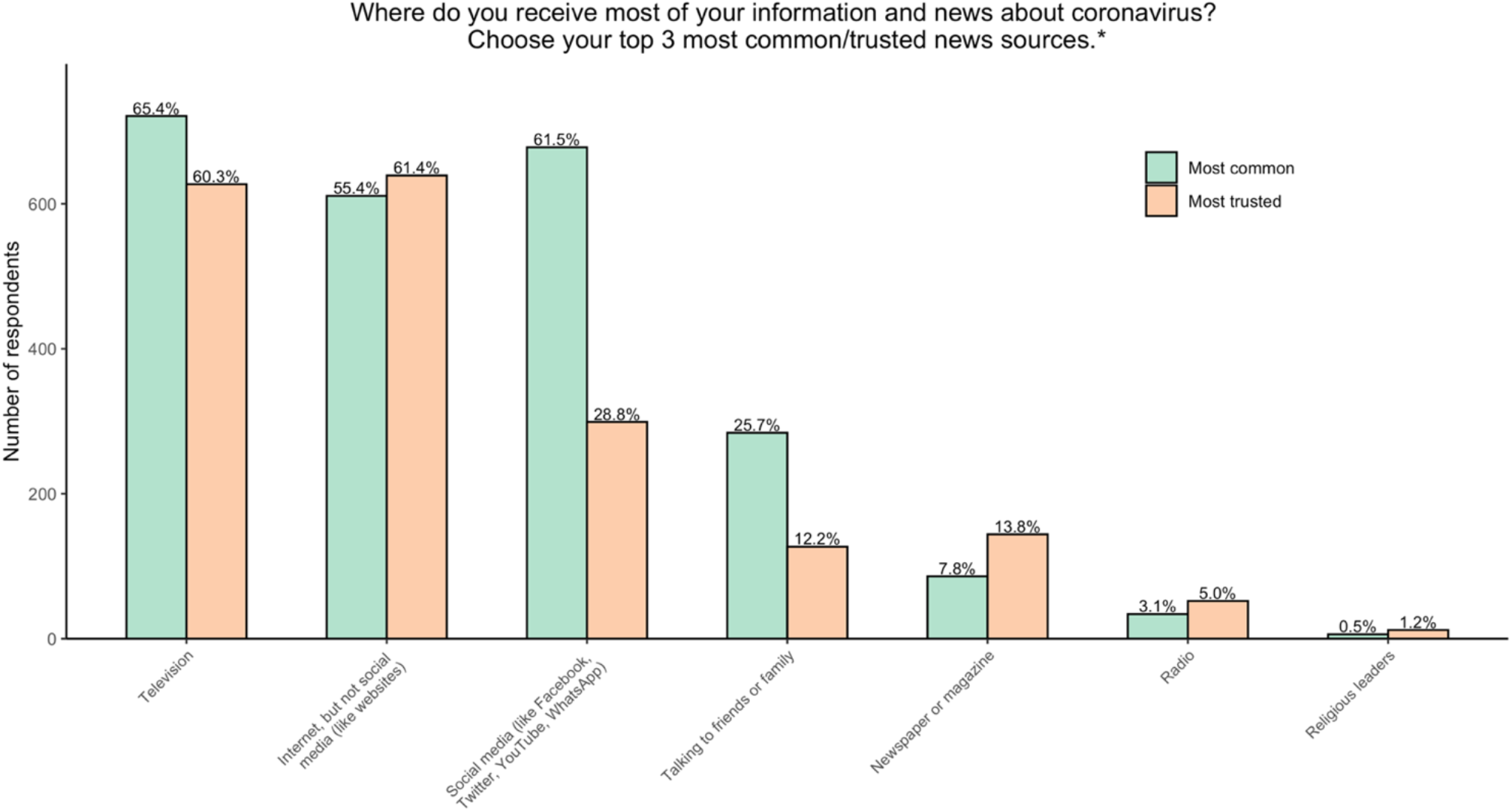
Most common and most trusted news sources about coronavirus. This figure summarizes responses to two separate questions, the first asking which news sources participants most commonly used, and the second asking which news sources they trusted most. Percentages of each response were calculated by dividing the number of participants who selected a response by the total number of participants who answered that question. ^*^The survey allowed participants to choose multiple answers for these questions; consequently, the sum of all subcategories does not equal the number of all participants who answered the questions.

### Qualitative Analysis

#### Top motivations for intending to vaccinate

Most frequently, participants cited the following motivations for their decisions to vaccinate: to protect themselves, their families, and the public, and to end the pandemic and return to normal life (Table 4). Somewhat frequently given reasons for intending to vaccinate included that participants felt like they “had no other choice” given the state of the pandemic, and that participants trusted science and research.

**Table 4:**
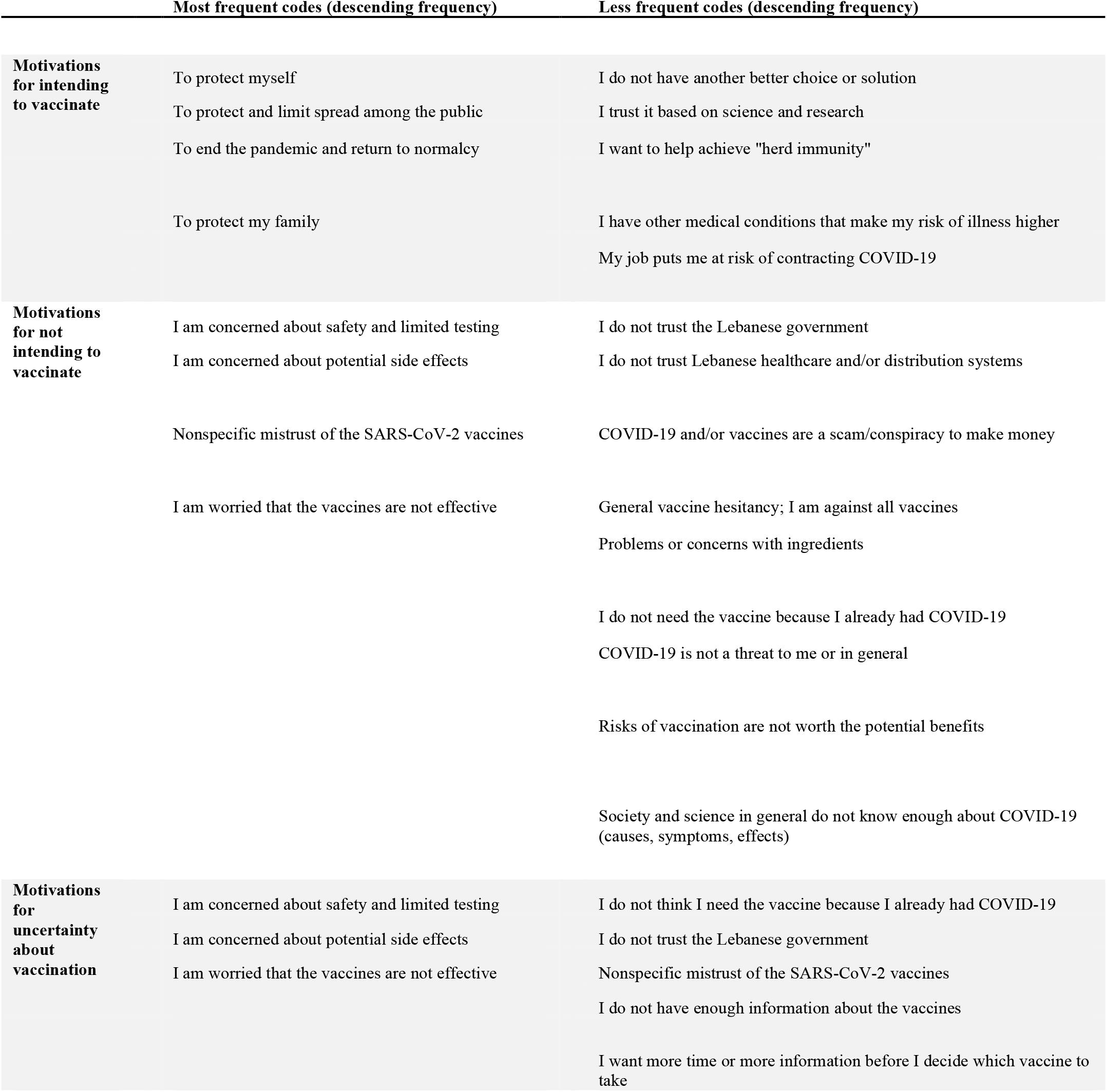
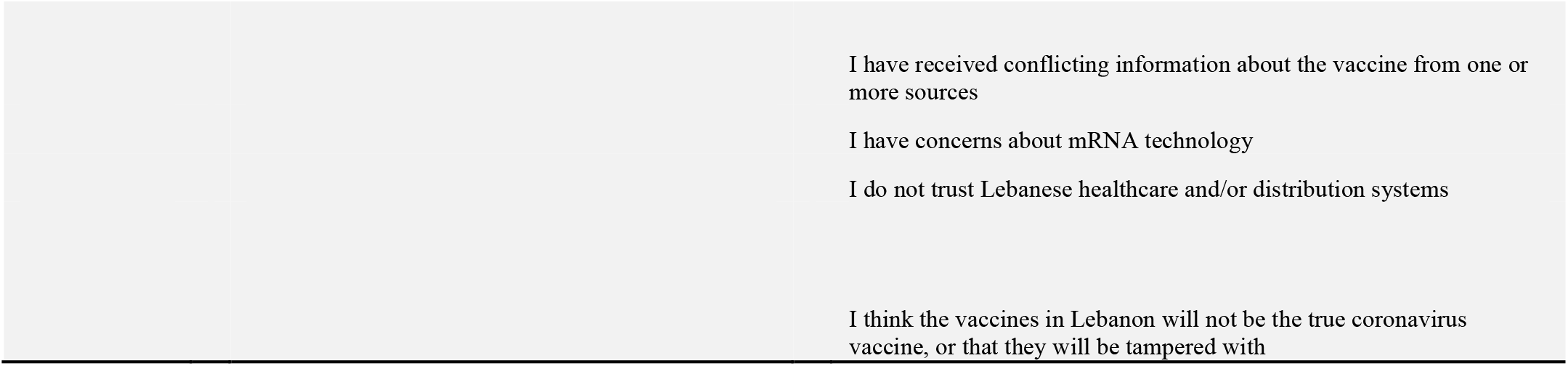
Summary of coded open-ended responses describing motivations for intentions about vaccination. This shows the most frequently observed codes in the optional open-ended responses to questions asking about motivations behind deciding to vaccinate, deciding not to vaccinate, and being uncertain about vaccination. For each group (intending to vaccinate, not intending to vaccinate, and uncertainty about vaccination), the denominator was the total number of participants in the group who provided a free-response to the question. ^a^”Most frequent codes” were observed in ≥15% of responses to the question. ^b^”Less frequent codes” were observed in 3%-15% of responses to the question.

| TABLE 4: SUMMARY OF CODED OPEN-ENDED RESPONSES DESCRIBING MOTIVATIONS |  |  |
| --- | --- | --- |
|  | Most frequent codes (descending frequency) | Less frequent codes (descending frequency) |
| **Motivations for intending to vaccinate** | To protect myself | I do not have another better choice or solution |
|  | To protect and limit spread among the public | I trust it based on science and research |
|  | To end the pandemic and return to normalcy | I want to help achieve "herd immunity" |
|  | To protect my family | I have other medical conditions that make my risk of illness higher<br>My job puts me at risk of contracting COVID-19 |
| **Motivations for not intending to vaccinate** | I am concerned about safety and limited testing | I do not trust the Lebanese government |
|  | I am concerned about potential side effects | I do not trust Lebanese healthcare and/or distribution systems |
|  | Nonspecific mistrust of the SARS-CoV-2 vaccines | COVID-19 and/or vaccines are a scam/conspiracy to make money |
|  | I am worried that the vaccines are not effective | General vaccine hesitancy; I am against all vaccines<br>Problems or concerns with ingredients |
|  |  | I do not need the vaccine because I already had COVID-19<br>COVID-19 is not a threat to me or in general |
|  |  | Risks of vaccination are not worth the potential benefits |
|  |  | Society and science in general do not know enough about COVID-19 (causes, symptoms, effects) |
| **Motivations for uncertainty about vaccination** | I am concerned about safety and limited testing | I do not think I need the vaccine because I already had COVID-19 |
|  | I am concerned about potential side effects | I do not trust the Lebanese government |
|  | I am worried that the vaccines are not effective | Nonspecific mistrust of the SARS-CoV-2 vaccines<br>I do not have enough information about the vaccines |
|  |  | I want more time or more information before I decide which vaccine to take |

#### Top reasons for vaccine hesitancy

Among participants who responded that they did not intend to be vaccinated, several themes emerged (Table 4). The most frequent reasons for vaccine hesitancy were concerns about safety given the fast development and limited testing of the vaccines, fears about side effects, and doubts about efficacy. Somewhat frequently cited concerns leading to vaccine hesitancy were mistrust in the Lebanese government.

#### Top reasons for uncertainty about vaccination

The most commonly provided reasons for uncertainty about whether participants planned to take the vaccine were similar: concerns about safety given the fast development and limited testing of the vaccines, fears about side effects, and doubts about efficacy (Table 4). Somewhat frequently mentioned concerns included mistrust of the Lebanese government and healthcare system, potential fraud in storage or marketing of the vaccines, wanting more information about the vaccines, and needing more time to decide which vaccine to take.

## DISCUSSION

One of our study’s primary objectives was to determine the proportion of Lebanese adult residents who intended to take (*vaccine acceptance*), who did not intend to take (*vaccine hesitancy*), and who were unsure about taking the SARS-CoV-2 vaccine (*vaccine uncertainty*) when available to them (while deciding to use these terms, we acknowledge that they imperfectly capture the complexity of individuals’ decisions about vaccines) [29]. Our findings of rates of vaccine acceptance and vaccine hesitancy were similar to studies of other countries in the Middle East and across the world. Vaccine acceptance of 46.1% in our sample was similar to rates in Kuwait (53.1%) and Qatar (45%-60%); higher than rates in Jordan (28.4%); and somewhat lower than in Saudi Arabia (64.7%) [23,30–32]. Rates of vaccine acceptance in our study were also similar to a large survey of 15 developed countries across the globe, in which 54% of respondents indicated they intend to vaccinate [33]. Other studies found higher rates of vaccine acceptance in specific developed countries—US (67%), Japan (62.1%), Ireland (65%), and the UK (69%) [34–36]. However, these studies were conducted earlier than our study, prior to initiation of mass-scale vaccination, and as systematic reviews of vaccine perception studies showed, vaccine acceptance and hesitancy have varied with time throughout the pandemic, with a trend toward decreasing acceptance throughout 2020 [20,37]. Indeed, our results show lower rates of vaccine acceptance than a survey in April-May 2020 that included a question about acceptance of hypothetical SARS-CoV-2 vaccines, in which 69.3% of Lebanese residents stated they would be willing to take a vaccine; it is important to note that at this time, no vaccines were developed or approved, and this survey’s population was 73.7% male and 89.0% aged younger than 45 years old, compared to our population which was 62.1% female and 79.1% aged younger than 45 years old [11].

One of our important secondary objectives was to assess which demographic characteristics were associated with vaccine hesitancy. Our findings of trends toward increased vaccine hesitancy in women, younger age groups, unemployed individuals, and individuals with lower education attainment are generally consistent with findings in the Middle East and globally, with the notable exception that in Kuwait and Qatar, hesitancy was increased in older populations [30,31,35–37]. While our study attempted to assess the association of vaccine hesitancy with religion and income, two important demographic variables given Lebanon’s social, political, and economic context, 15.9% and 36.3% of participants skipped the questions about religion and income, respectively, demonstrating the topics’ sensitive natures. Therefore, we do not recommend making inferences from our study about differences in vaccine acceptance or hesitancy by religion or income. Similarly, refugees (n = 56, 5.1%) were underrepresented in our sample. While no significant difference in vaccine acceptance by refugee status emerged, further focus on vaccination in refugees in Lebanon is merited given their multiple vulnerabilities.

Another important secondary objective of our study was to evaluate how individuals’ experiences with COVID-19 affected vaccine acceptance. Unsurprisingly, frequent mask wearing was associated with increased vaccine acceptance, likely because these individuals see COVID-19 as a more serious threat than those who do not wear masks frequently. The non-significant trend toward increased vaccine acceptance among participants with a close acquaintance who contracted COVID-19 could imply that personal experience with someone affected by the virus increases willingness to vaccinate. Interestingly, however, neither personal history of infection nor history of infection in a close contact was consistently associated with vaccine acceptance in a meta-analysis of SARS-CoV-2 perception studies [37].

The timing of our survey period spanned the initiation of vaccination in Lebanon. While our data suggested increased vaccine acceptance among participants who completed the survey after initiation of vaccination, this must be interpreted cautiously. The demographics of the respondents after initiation of vaccination were significantly different from those who completed the survey before initiation of vaccination. We believe that the differences in vaccine acceptance before and after initiation of vaccination more likely reflect differences in populations surveyed during these periods, given the selection bias inherent in the convenience “snowball” sampling method.

Our study was conceived with the goal of providing information that would be useful for implementation of vaccination efforts in Lebanon. The logistical considerations about which we asked can provide some guidance. The most commonly selected sources of news about COVID-19 were television, social media, and internet websites; among these, television and internet websites were the most trusted. Though religion is influential in Lebanon, only 6 (0.5%) participants cited religious leaders as an important source of COVD-19 news. Given that respondents who selected newspapers, magazines, and radio were more likely to intend to vaccinate, focusing dissemination of vaccine promotion efforts on television, social media, and internet websites would appear to be most efficient to reach those who are hesitant to vaccinate.

Survey respondents also reported preferring to receive the vaccine at familiar, established health care sites: hospitals, doctors’ offices, primary health centers, and pharmacies. While temporary dedicated vaccination centers were not popular, a small but significant number of participants (106, 11.8%) stated they would prefer vaccination by a visiting medical professional to their homes. This could be an important means of reaching vulnerable patients willing to be vaccinated but unable to go to vaccination sites. A final logistical consideration about which our survey asked was whether a financial incentive would change participants’ minds so that they decide to vaccinate; overwhelmingly, they indicated that it would not (98.3%). Based on this study, it appears that offering a financial incentive would not be an efficient means of increasing vaccination rates in Lebanon.

Perhaps the most actionable information generated in this study involves motivations for vaccine acceptance and vaccine hesitancy. The most frequently provided reasons for intending to vaccinate were to protect oneself, to protect one’s family and the public, and to end the pandemic. These goals are similar to the most common motivations in the Middle East and globally [30,31,37]. In our study, the most commonly provided reasons for SARS-CoV-2 vaccine hesitancy involved concerns about its safety given the fast development and limited testing of the vaccines (including regarding the mRNA technology), fears about side effects, and doubts about efficacy. Somewhat frequently cited concerns leading to vaccine hesitancy were mistrust in the Lebanese government. Other reasons mentioned for vaccine uncertainty included worrying about potential fraud in storage or marketing of the vaccines, wanting more information about the vaccines, and needing more time to decide which vaccine to take. These results are consistent with those of studies globally [37,38]. While several participants in our study cited conspiracy theories as reasons for not vaccinating, these were relatively uncommon, especially compared to a study in other Arabic speaking countries, which found rates of belief in conspiracy theories of over 50% [32].

Interestingly, logistical factors were not frequently of concern among adults living in Lebanon. Despite financial hardships in Lebanon, barriers to vaccine access (cost, transportation, proximity to medical care) were not cited frequently as concerns about vaccination in our study. This might be explained by the fact that it has become common for governments, including in developing countries, to distribute the vaccine free of charge as a part of public health mandates. While a few participants did express preference for “the vaccine from China or Russia,” most perceptions applied generally to all SARS-CoV-2 vaccines, and there were relatively few concerns about vaccine properties like number of required doses, country of origin, or specific vaccine brands. Also uncommon in our study was generalized opposition to vaccination, i.e., not specific to SARS-CoV-2 vaccines. This is consistent with previous studies in Lebanon showing moderate uptake of routine vaccinations [12,14,15,17,39].

The relatively large proportion of participants in our study who were uncertain about vaccination—34.0%—provides a public health opportunity and imperative in the effort to achieve mass vaccination in Lebanon. The majority of concerns about SARS-CoV-2 vaccination involved absence of reliable information and data of safety and efficacy for this new medical technology. Increasing public availability in Lebanon of high-quality, data-driven, up to date, accessible information about the SARS-CoV-2 vaccines could assuage some of these concerns and increase vaccine acceptance. Based on our results about common COVID-19 news sources, we recommend disseminating clear, consistent, verifiable safety and efficacy information on television, social media, and news websites. Given the prevalence of mistrust in the government, third parties (like healthcare organizations) might be most likely to be trusted, especially if they promote vaccination using the motivators that those in our sample cited as reasons for vaccination.

Our study has several strengths. It is, to our knowledge, the first such academic study about SARS-CoV-2 vaccine intentions and perceptions in Lebanon. Its online platform allowed for rapid surveying of a large sample size remotely. The open-ended nature of the questions about reasons why participants intended or did not intend to vaccinate, or were uncertain about vaccination, allowed for participants to share their most central motivations without suggestive multiple-choice options, producing what we believe is a more authentic understanding of motivations and concerns.

There are several important limitations of our study. First and foremost, our sample is unlikely to be representative of the general population because of the sampling strategy. While we attempted to mitigate this by collecting and describing important demographic and experiential characteristics, the bias remains, and several important demographic groups were underrepresented, most notably the elderly, members of Shi’a and Sunni religions, residents outside of Beirut and Mount Lebanon, non-Lebanese citizens, and those who identify as refugees. Small sample sizes of subgroups limited analysis of associations with certain variables, especially governorate and citizenship. Second, participation required literacy in Arabic, access to the internet, and digital literacy, potentially excluding some populations (though it is noteworthy that >80% of refugees in Lebanon have access to mobile technology like WhatsApp) [40]. Third, some participants did not answer all questions, possibly leading to non-response bias. Finally, vaccine hesitancy, perceptions, and concerns may be changing rapidly over time; our results should be interpreted as pertaining to the time period during which the survey was conducted.

## CONCLUSION

This cross-sectional study assessed intentions to vaccinate against SARS-CoV-2 among adults residing in Lebanon, analyzed characteristics that were associated with vaccine acceptance and hesitancy, and described motivations for and concerns about vaccination. We recommend disseminating clear, consistent, evidence-based safety and efficacy information on vaccines on the most commonly reported news sources by participants: television, social media, and news websites. As vaccination efforts continue, repeated assessments of intentions to vaccinate, concerns or obstacles regarding vaccination, and changes in motivations should be performed, especially with the goal of assessing the perspectives and needs of populations that were underrepresented in this study.

## Supporting information

Qualitative Response Coding Protocol

Recruitment Script Arabic

Recruitment Script English

Survey Arabic

Survey English

Tables and Figures included in main manuscript

## Data Availability

Deidentified data from this study is available upon request from and will be stored in the appendix of the corresponding scientific journal in which it is eventually published.

## ETHICAL APPROVAL AND INFORMED CONSENT

This study was approved by the Institutional Review Boards of the University of California, Berkeley; the Modern University for Business and Science, Lebanon; and Stanford University (by reliance agreement with the University of California, Berkeley). Informed consent was obtained from all participants prior to the survey; a waiver of written informed consent was granted.

## ACKNOWLEDGEMENTS

We thank our participants for taking the time to share their perspectives; our recruitment and coding assistants (Hanin Abi Ghannam, Jana Al Khatib, Nathalie Al Saady, Ghiwa Nasr) for disseminating the survey and synthesizing our data; the National Wellness Network and Mariam Fadel for assisting in disseminating the survey; the Stanford King Center on Global Development for awarding us a Capacity Building and Policy Engagement Grant; the Stanford Center for Innovation in Global Health and the Refugee and Asylum seeker Health Initiative (RAHI) at the University of California, San Francisco for their cultivation of international collaborations and fostering global health equity; and Drs. Michele Barry, Cybele Renault, Karen Sokal-Gutierrez, and Anke Hemmerling for their mentorship.

