## Supplementary material for "Perceptions of and obstacles to SARS-CoV-2 vaccination among adults in Lebanon: a cross-sectional online survey": Qualitative Response Coding Protocol

**Process**

**Step 1**: *Nadeem*

**Create an entry** for each participant in our qualitative analysis data spreadsheet by filling in each column in the “Data” tab of the spreadsheet.

1) Subject ID (copied and pasted from survey response raw data)
2) Arabic responses to questions (copied and pasted from survey response raw data by Nadeem)

**Step 2**: *Jana, Ghiwa, Hanin, Nathalie, Roua, Vanessa*

**Translate Arabic responses into English.**

When the primary translator has translated a row, please change the color of the corresponding box in the "Translators" column to this color orange.
When the verifying translator has translated a row, please change the color of the corresponding box in the "Translators" column to this color green.

If you are verifying the translation and believe it should be changed, please contact the initial translator and discuss. If you cannot agree upon the translation, please contact Diana for help.

1) For subjects in rows 1-50, Jana will do the initial translation and Vanessa will verify it
2) For subjects in rows 51-100, Nathalie will do the initial translation and Jana will verify it
3) For subjects in rows 101-150, Hanin will do the initial translation and Nathalie will verify it
4) For subjects in rows 151-200, Roua will do the initial translation and Hanin will verify it
5) For subjects in rows 201-250, Ghiwa will do the initial translation and Roua will verify it
6) For subjects in rows 251-300, Vanessa will do the initial translation and Ghiwa will verify
7) For subjects in rows 301-350, Jana will do the initial translation and Vanessa will verify it
8) For subjects in rows 351-400, Nathalie will do the initial translation and Jana will verify it
9) For subjects in rows 401-450, Hanin will do the initial translation and Nathalie will verify it
10) For subjects in rows 451-500, Roua will do the initial translation and Hanin will verify it
11) For subjects in rows 501-550, Ghiwa will do the initial translation and Roua will verify it
12) For subjects in rows 551-600, Vanessa will do the initial translation and Ghiwa will verify
13) For subjects in rows 601-650, Jana will do the initial translation and Vanessa will verify it
14) For subjects in rows 651-700, Nathalie will do the initial translation and Jana will verify it
15) For subjects in rows 701-750, Hanin will do the initial translation and Nathalie will verify it
16) For subjects in rows 751-800, Roua will do the initial translation and Hanin will verify it
17) For subjects in rows 801-850, Ghiwa will do the initial translation and Roua will verify it
18) For subjects in rows 851-900, Vanessa will do the initial translation and Ghiwa will verify
19) For subjects in rows 901-950, Jana will do the initial translation and Vanessa will verify it
20) For subjects in rows 951-967, Nathalie will do the initial translation and Jana will verify
21) For subjects in rows 968-1000, Roua will do the initial translation, Vanessa will verify
22) For subjects in rows 1001-1100, Vanessa will do the initial translation, Roua will verify
23) For subjects in rows 1101-1200, Roua will do the initial translation, Vanessa will verify
24) For subjects in rows 1201-1300, Vanessa will do the initial translation, Roua will verify

25) For subjects in rows 1301-1400, Roua will do the initial translation, Vanessa will verify

**Step 3**: *Jana, Ghiwa, Hanin, Nadeem, Nadine, Nathalie, Roua, Vanessa*
**Assign qualitative codes to responses.**

Using the codes in the “Codes” tab of the qualitative analysis data spreadsheet, apply all relevant codes to the free response questions for your assigned subjects. If you think a new code needs to be created, please create one (see “Notes” section below for more information).

When the primary coder has finished coding for a row, please change the color of the corresponding box in the "Coders" column to this color orange.
When the verifying coder has finished coding for a row, please change the color of the corresponding box in the "Coders" column to this color green.

If you are verifying the coding and believe it should be changed, please contact the initial coder and discuss. If you cannot agree upon the coding, please contact Nadeem and Diana for help. To ensure consistency, Nadeem will review all coding.

1) For subjects in rows 1-50, Jana will do the initial coding and Nadeem will verify it
2) For subjects in rows 51-100, Nadeem will do the initial coding and Nathalie will verify it
3) For subjects in rows 101-150, Nathalie will do the initial coding and Nadine will verify it
4) For subjects in rows 151-200, Nadine will do the initial coding and Hanin will verify it
5) For subjects in rows 201-250, Hanin will do the initial coding and Roua will verify it
6) For subjects in rows 251-300, Roua will do the initial coding and Ghiwa will verify it
7) For subjects in rows 301-350, Ghiwa will do the initial coding and Vanessa will verify it
8) For subjects in rows 351-400, Vanessa will do the initial coding and Jana will verify it
9) For subjects in rows 401-450, Jana will do the initial coding and Nadeem will verify it
10) For subjects in rows 451-500, Nadeem will do the initial coding and Nathalie will verify it
11) For subjects in rows 501-550, Nathalie will do the initial coding and Nadine will verify it
12) For subjects in rows 551-600, Nadine will do the initial coding and Hanin will verify it
13) For subjects in rows 601-650, Hanin will do the initial coding and Roua will verify it
14) For subjects in rows 651-700, Roua will do the initial coding and Ghiwa will verify it
15) For subjects in rows 701-750, Ghiwa will do the initial coding and Vanessa will verify it
16) For subjects in rows 751-800, Vanessa will do the initial coding and Jana will verify it
17) For subjects in rows 801-850, Jana will do the initial coding and Nadeem will verify it
18) For subjects in rows 851-900, Nadeem will do the initial coding and Nathalie will verify it
19) For subjects in rows 901-950, Nathalie will do the initial coding and Nadine will verify it
20) For subjects in rows 951-967, Nadine will do the initial coding and Hanin will verify it
21) For subjects in rows 968-1000, Nadeem will do the initial coding and Vanessa will verify
22) For subjects in rows 1001-1050, Roua will do the initial coding and Nadeem will verify it
23) For subjects in rows 1051-1100, Nadine will do the initial coding and Roua will verify it
24) For subjects in rows 1101-1150, Vanessa will do the initial coding and Nadine will verify
25) For subjects in rows 1151-1200, Nadeem will do the initial coding and Vanessa will verify
26) For subjects in rows 1201-1250, Roua will do the initial coding and Nadeem will verify it

27) For subjects in rows 1251-1300, Nadine will do the initial coding and Roua will verify it
28) For subjects in rows 1301-1350, Vanessa will do the initial coding and Nadine will verify
29) For subjects in rows 1351-1400, Nadeem will do the initial coding and Vanessa will verify

**Step 4:** *Nadeem*
**Ensure consistency between codes**

Nadeem will review all codes and responses to assure that codes are being used and applied consistently between all responses.

**Notes**
General
- The Qualitative Analysis Data Spreadsheet is an edited version of the full survey responses. It only includes the Response ID, which is a code that was randomly assigned to each participant by the survey software, and the survey participant's responses to the qualitative (free-response) questions
- Each row is an individual survey participant's responses
- DO NOT edit the ResponseID column. It is very important that this column remains the same
- If they already replied in English, please simply copy and paste their English reply into the "English" column.
- There will be many empty data cells. This is ok. Empty data cells exist because participants only were asked one question from the categories Q6-Q8, and Q15-Q17. Participants were not required to answer any of these questions.

Coding
- If you would like to reference the questions that correspond to the responses, please open the second tab on the spreadsheet. Sometimes referencing these questions can help you code a response more correctly.
- The third tab of the spreadsheet has the list of codes that we will be applying to the responses. Please use the abbreviation that is listed for the descriptive code that you believe applies
- Please include all codes that apply to the response that you are coding, separated by commas
- If you cannot find a code that applies to a response, you can create a new one. Please create both 1) a descriptive code and 2) a four-letter code abbreviation that is unique and has not been used by another code.
- If you have a question about how to code, please see the example in the fourth tab of the spreadsheet
- If you and your co-coder cannot agree on appropriate coding, please ask Nadeem or Vanessa for assistance
