## Supplementary material for "Perceptions of and obstacles to SARS-CoV-2 vaccination among adults in Lebanon: a cross-sectional online survey": Recruitment Script Arabic

مرحباً. هذه رسالة من الدكتور أبو عرّاج من جامعتيّ ستانفورد (Stanford) و بيركلي (Berkeley) في الولايات المتحدة الأمريكية و الدكتور علامي من الجامعة الحديثة للإدارة والعلوم (MUBS) في لبنان.

نجري دراسة بعنوان "شو رأيك؟" لأخذ رأيك حول لقاح فيروس كورونا.

هل يمكنك مساعدتنا من خلال المشاركة في هذا الإستطلاع القصير؟ مشاركتك طوعية تماماً.

للمشاركة، يجب أن يكون عمرك ١٨ عاماً أو أكثر ويجب أن تقيم حالياً في لبنان. لن يتم ربط رقم هاتفك بأجوبتك. لن يجمع الاستطلاع إسمك أو أي معلومات أُخرى يمكن أن تحدد هويتك.

نتمنى تجميع الآراء من جميع أنحاء لبنان. لذلك يرجى ارسال هذه الرسالة التي تتضمن رابط الاستطلاع إلى اصدقائك الراشدين و عائلتك في لبنان، وخاصة كبار السن او الذين ليس لهم وسيلة لاستخدام مواقع التواصل الاجتماعي.

يمكن لكل جهاز محمول الدخول الى الاستبيان مرة واحدة فقط، لذلك يجب على كل مشارك جديد أن يقوم بتعبئة الاستبيان على جهازه الخاص.

إذا كنت ترغب في مساعدتنا فيرجى الضغط على رابط الإستطلاع أدناه:

https://berkeley.qualtrics.com/jfe/form/SV_eOJ20K6h2wy5jfw
