## Supplementary material for "Perceptions of and obstacles to SARS-CoV-2 vaccination among adults in Lebanon: a cross-sectional online survey": Recruitment Script English

Hello. This is Dr. Abou-Arraj from Stanford and Berkeley universities in the USA and Dr. Alami from the Modern University for Business and Science (MUBS) in Lebanon. We are conducting a research study called "Shou Raa'yak?" to understand perceptions of coronavirus vaccination.

Could you please help us by taking this short, anonymous survey?

To participate, you must be 18 years or older and live in Lebanon. Your phone number will not be linked to your responses. We will not collect any information that can identify you.

We hope to gather opinions from all over Lebanon. Please forward this message and survey to your adult friends and family in Lebanon, especially those who are elderly or do not have easy access to social media.

Each mobile device can only access the survey once, so each new participant must take the survey on a different device.

If you would like to help us, please click this link to access the survey:
[Survey link]
