## Supplementary material for "Perceptions of and obstacles to SARS-CoV-2 vaccination among adults in Lebanon: a cross-sectional online survey": Survey Arabic

**الموافقة و التعليمات:**

شكراً لك على اهتمامك ب "شو رايك؟"، استطلاع حول فيروس كورونا و لقاح لفيروس كورونا.

ستساعد هذه الدراسة الباحثين من الجامعة الحديثة للادارة والعلوم في بيروت وجامعة كاليفورنيا، بيركلي وجامعة ستانفورد في الولايات المتحدة على مساعدة لبنان في مواجهة فيروس كورونا.

هذا الاستطلاع لا يجمع إسمك أو أي معلومات يمكن أن تحدد هويتك. نظرًا بأن هذه دراسة بحثية، فسوف نعلمك بالفوائد والمخاطر المحتملة للمشاركة، والتي تم وصفها في السؤال رقم ١. لن يتم تعويضك عن مشاركتك.

إذا كنت تعاني من أعراض أو مخاوف صحية اخرى، فيجب عليك طلب الإستشارة من مقدّم الرعاية الصحية/طبيب. إذا كنت تعاني من حالة طارئة، فيرجى إيقاف هذا الاستطلاع و العثور على طبيب او مستشفى، او الاتصال على ٠١٥٩٤٤٥٩ الخط الساخن الخاص بوزارة الصحة العامة لفيروس كورونا.

#١. يُرجى قراءة وثيقة الموافقة المفصّلة المتاحة أدناه. هل تؤكد أنك قد قرأت وثيقة الموافقة، و فهمت المخاطر و الفوائد، و ترغب بالمشاركة في الدراسة البحثية؟

**[سيتم وضع صورة لوثيقة الموافقة هنا]**

١. اجل، افهم و ارغب بالمشاركة في الدراسة البحثية.

٢. لا أرغب بالمشاركة في الدراسة البحثية.

**التركيبة السكانية الأساسية:**

#٢. كم تبلغ من العمر (بالسنوات)؟ الرجاء التحديد من هذه القائمة أدناه.

#٣. هل تعيش في لبنان حالياً؟

١. نعم

٢. لا

**لقاح فيروس كورونا:**

#٤. الاسئلة التالية تتعلق بالصحة وفيروس كورونا (يشار الى المرض ايضاً باسم "كوفيد ١٩"، الناجم عن الفيروس الجديد "SARS-CoV-2").

هل سمعت عن فيروس كورونا؟

١. نعم

٢. لا

#٥. طوّر العلماء لقاحات لحماية الأفراد من فيروس كورونا.
في حال أصبح اللقاح متاحاً لك، هل تخطط للحصول عليه؟

اختر افضل اجابة.

١. نعم

٢. لا

٣. غير متأكد

٤. تخطي هذا السؤال

#٦. لماذا تخطط للحصول على لقاح فيروس كورونا في حال أصبح متاحاً لك؟

يرجى صياغة جوابك في بضع جمل. اذا كنت لا ترغب في الاجابة، اترك الاجابة فارغة و انتقل الى السؤال التالي.

#٧. لماذا **لا تريد** الحصول على لقاح فيروس كورونا في حال أصبح متاحاً لك؟

يرجى صياغة جوابك في بضع جمل. اذا كنت لا ترغب في الاجابة، اترك الاجابة فارغة و انتقل الى السؤال التالي.

#٨. لماذا انت **غير متأكد** من حصولك على لقاح فيروس كورونا في حال أصبح متاحاً لك؟

يرجى صياغة جوابك في بضع جمل. اذا كنت لا ترغب في الاجابة، اترك الاجابة فارغة و انتقل الى السؤال التالي.

#٩. للتوضيح، **لن** يكون هناك أي مكافأة مالية لأحد من أجل الحصول على لقاح فيروس كورونا. ومع ذلك، **افترض** أنه عُرض عليك المال، فهل سيغيّر ذلك رأيك حتى تقرر الحصول على لقاح فيروس كورونا؟

١. نعم

٢. لا

٣. تخطي هذا السؤال

#١٠. ما هو المبلغ المالي الذي تحتاجه من أجل تغيير رأيك للحصول على لقاح فيروس كورونا؟ يرجى تحديد **جميع** المبالغ المالية التي ستكون كافية لتغيير رأيك حتى تحصل على لقاح فيروس كورونا.

على سبيل المثال، **افترض** أنك ستأخذ اللقاح إذا تلقيت ٢٥،٠٠٠ ليرة لبنانية ، لكنك **لن** تأخذ اللقاح إذا عُرض عليك **أقل** من ذلك. في هذه الحالة، سوف تختار الإجابات ١ و ٢ و ٣ و ٤.

مرة أخرى، للتوضيح، **لن** يكون هناك أي مكافأة مالية لأحد من أجل الحصول على لقاح فيروس كورونا. هذا سؤال **افتراضي** فقط.

إذا كنت لا ترغب في الإجابة ، يُرجى اختيار "تخطي هذا السؤال".

[اختر كل ما ينطبق]

١. أكثر من ١٠٠،٠٠٠ ل.ل.

٢. بين ٧٦،٠٠٠ ل.ل. و ١٠٠،٠٠٠ ل.ل.

٣. بين ٥١،٠٠٠ ل.ل. و ٧٥،٠٠٠ ل.ل.

٤. بين ٢١،٠٠٠ ل.ل. و ٥٠،٠٠٠ ل.ل.

٥. بين ١١،٠٠٠ ل.ل. و ٢٠،٠٠٠ ل.ل.

٦. بين ١،٠٠٠ ل.ل. و ١٠،٠٠٠ ل.ل.

٧. **لن** احصل على لقاح فيروس كورونا حتى لو كان مجاناً.

٨. تخطي هذا السؤال.

#١١. للتوضيح، **لم** يتم التحديد إذا سيكون لقاح فيروس كورونا مجانياً للأفراد الذين سيتم تطعيمهم أو إذا سيكلف المال. ومع ذلك، يرجى **الإفتراض** أنه يمكنك شراء لقاح. ما **أقصى** مبلغ ترغب في دفعه من أموالك (لا يشمل التأمين) مقابل لقاح فيروس كورونا عندما يصبح متاحاً؟

إختر أفضل إجابة.

١. صفر ل.ل.

٢. بين ١،٠٠٠ ل.ل. و ١٠،٠٠٠ ل.ل.

٣. بين ١١،٠٠٠ ل.ل. و ٢٠،٠٠٠ ل.ل.

٤. بين ٢١،٠٠٠ ل.ل. و ٥٠،٠٠٠ ل.ل.

٥. بين ٥١،٠٠٠ ل.ل. و ٧٥،٠٠٠ ل.ل.

٦. بين ٧٦،٠٠٠ ل.ل. و ١٠٠،٠٠٠ ل.ل.

٧. أكثر من ١٠٠،٠٠٠ ل.ل.

٨. **لن** احصل على لقاح فيروس كورونا حتى لو كان مجاناً.

٩. تخطي هذا السؤال.

#١٢. أين تفضل الحصول على لقاح فيروس كورونا؟

إختر أفضل ٣ مواقع بالنسبة لك.

١. عيادة طبيبي

٢. أي عيادة طبيب

٣. مركز صحي أوَّلي/مستوصف

٤. صيدلية

٥. مستشفى

٦. مبنى قريب من منزلي يُستخدم كعيادة تطعيم مؤقتة

٧. شاحنة متنقلة قريبة من منزلي تعمل كعيادة تطعيم مؤقتة

٨. في منزلي، تقام من قبل مقدم تطعيم معتمد

٩. إجابة أخرى

١٠. **لن** احصل على لقاح فيروس كورونا بغض النظر عن المواقع المتاحة

١١. تخطي هذا السؤال

#١٣. يرجى وصف المكان الذي تفضل تلقي لقاح فيروس كورونا فيه.

يرجى صياغة جوابك في بضع جمل. اذا كنت لا ترغب في الاجابة، اترك الاجابة فارغة و انتقل الى السؤال التالي.

#١٤. اذا كان لديك أطفال دون سن ١٨ ، فهل تخطط أن يتلقى اطفالك لقاح فيروس كورونا إذا أصبح متاحاً لك و أوصى به الاطباء من اجل الاطفال؟

إختر افضل اجابة.

١. نعم

٢. لا

٣. لست متاكداً

٤. ليس لدي اطفال

٥. تخطي هذا السؤال

#١٥. لماذا تريد أن يتلقى اطفالك لقاح فيروس كورونا عندما يصبح متاحاً لك و موصى به للاطفال؟

يرجى صياغة جوابك في بضع جمل. اذا كنت لا ترغب في الاجابة، اترك الاجابة فارغة و انتقل الى السؤال التالي.

#١٦. لماذا **لا** تريد أن يتلقى اطفالك لقاح فيروس كورونا عندما يصبح متوفراً و موصى به للاطفال؟

يرجى صياغة جوابك في بضع جمل. إذا كنت لا ترغب في الاجابة، اترك الاجابة فارغة و انتقل الى السؤال التالي.

#١٧. لماذا انت **غير متأكد** من تلقي أطفالك لقاح فيروس كورونا عندما يصبح متوفراً و موصى به للاطفال؟

يرجى صياغة جوابك في بضع جمل. إذا كنت لا ترغب في الاجابة، اترك الاجابة فارغة و انتقل الى السؤال التالي.

**تجربة فيروس كورونا**

#١٨. حسب مفهومك الشخصي، كيف يصاب شخص ما بفيروس كورونا؟

إختر افضل اجابة.

١. تناول الطعام النيئ أو شرب المياه غير المكررة

٢. أن تكون قريباً جسدياً من شخص مصاب بفيروس كورونا

٣. لدغة حشرة

٤. تخطي هذا السؤال

#١٩. هل سبق أن تم تشخيصك بفيروس كورونا، أو كنت تعتقد أنك مصاب بفيروس كورونا بسبب أعراض صحية ولكن لم يتم تشخيصك من قبل طبيب؟

١. نعم

٢. لا

٣. تخطي هذا السؤال

#٢٠. هل تعرف شخصاً من عائلتك أو صديق مقرب لك مصاب أو أُصيب بفيروس كورونا؟

١. نعم

٢. لا

٣. تخطي هذا السؤال

#٢١. كم مرّة ترتدي قناع الوجه عندما تكون خارج منزلك؟

إختر أفضل إجابة.

١. أبداً

٢. نادراّ

٣. في بعض الأحيان

٤. معظم الوقت

٥. دائماً

٦. تخطي هذا السؤال

#٢٢. من أين تتلقى معظم معلوماتك وأخبارك حول فيروس كورونا؟

إختر مما يلي أكثر ٣ مصادر إخبارية **إستخداماً** بالنسبة لك.

١. جريدة أو مجلة

٢. راديو

٣. التلفزيون

٤. وسائل التواصل الاجتماعي، مثل "فيسبوك" و "تويتر" و "يوتيوب" و "واتساب"

٥. الإنترنت، ولكن ليس وسائل التواصل الاجتماعي، مثل موقع إلكتروني

٦. التحدث إلى الأصدقاء أو العائلة

٧. رجال الدين

٨. تخطي هذا السؤال

#٢٣. ما هو مصدر الأخبار الذي تثق به أكثر للحصول على معلومات وأخبار حول فيروس كورونا؟

إختر مما يلي أكثر ٣ مصادر إخبارية **موثوق بها** بالنسبة لك.

١. جريدة أو مجلة

٢. راديو

٣. التلفزيون

٤. وسائل التواصل الاجتماعي، مثل "فيسبوك" و "تويتر" و "يوتيوب" و "واتساب"

٥. الإنترنت، ولكن ليس وسائل التواصل الاجتماعي، مثل موقع إلكتروني

٦. التحدث إلى الأصدقاء أو العائلة

٧. رجال الدين

٨. تخطي هذا السؤال

**ديموغرافيات إضافية:**

٢٤#. اختر الكلمة التي تصفك بشكل أفضل وأدق.

إختر أفضل إجابة.

١. أنثى

٢. ذكر

٣. إجابة أُخرى

٤. تخطي هذا السؤال

٢٥#. في أي محافظة تسكن؟

إختر أفضل إجابة.

١. بعلبك - الهرمل

٢. البقاع

٣. بيروت

٤. جبل لبنان

٥. الجنوب

٦. عكار

٧. الشمال

٨. النبطية

٩. تخطي هذا السؤال

٢٦#. ما هو أعلى مستوى تعليمي أكملته؟

إختر أفضل إجابة.

١. لم أكمل دراستي/ لم اتعلم

٢. المدرسة الابتدائية

٣. المدرسة الثانوية

٤. الكلية أو الجامعة

٥. مدرسة مهنية

٦. مرحلة تخصص ما بعد الجامعة

٧. تخطي هذا السؤال

#٢٧. الى أي ديانة تنتمي؟

إختر أفضل إجابة.

١. تخطي هذا السؤال

٢. سنّة

٣. شيعة

٤. دروز

٥. مسلم آخر

٦. الموارنة

٧. الكاثوليكية

٨. الارثوذكسية

٩. مسيحي آخر

١٠. ديانات أخرى أو ديانات متعددة

١١. لا دين

#٢٨. ما هو وضعك الوظيفي؟

إختر أفضل إجابة.

١. موظف وأعمل حصريا من المنزل

٢. موظف وأذهب إلى العمل شخصياً لبعض الوقت أو بشكل دائم

٣. لا أعمل وأبحث عن عمل

٤. لا أعمل ولا أبحث عن عمل

٥. متقاعد

٦. رب/ربة منزل

٧. تلميذ

٨. تخطي هذا السؤال

#٢٩. ما هو إجمالي الدخل الأسري الخاص بك المقدر لعام ٢٠١٩؟

إختر أفضل إجابة.

١. تخطي هذا السؤال

٢. أقل من ١،٠٠٠،٠٠٠ ل.ل

٣. بين ١،٠٠٠،٠٠٠ ل.ل و ١،٩٩٩،٩٩٩ ل.ل

٤. بين ٢،٠٠٠،٠٠٠ ل.ل و ٩،٩٩٩،٩٩٩ ل.ل

٥. بين ١٠،٠٠٠،٠٠٠ ل.ل و ١٩،٩٩٩،٩٩٩ ل.ل

٦. بين ٢٠،٠٠٠،٠٠٠ ل.ل و ٣٩،٩٩٩،٩٩٩ ل.ل

٧. بين ٤٠،٠٠٠،٠٠٠ ل.ل. و ٦٩،٩٩٩،٩٩٩ ل.ل

٨. بين ٧٠،٠٠٠،٠٠٠ ل.ل و ٩٩،٩٩٩،٩٩٩ ل.ل

٩. ١٠٠،٠٠٠،٠٠٠ ل.ل. أو أكثر

٣٠#. ما هي جنسيتك؟ إذا كنت مواطنًا في أكثر من دولة، يمكنك اختيار أكثر من إجابة واحدة.

١. لبناني

٢. سوري

٣. فلسطيني

٤. دولة أخرى في الشرق الأوسط، مثل العراق، مصر، الأردن، وغيرها

٥. بلاد جنوب أفريقيا ما عدا البلاد العربية، مثل اثيوبيا، كينيا، وغيرها

٦. بلد في جنوب آسيا، مثل بنغلاديش، سريلانكا، الهند، وغيرها

٧. بلد جزر المحيط الهادئ، مثل الفلبين، إندونيسيا، وغيرها

٨. بلد في أوروبا أو أمريكا الشمالية، مثل فرنسا، كندا، وغيرها

٩. بلد آخر

١٠ . تخطي هذا السؤال

٣١#. هل أنت لاجئ مقيم في لبنان؟

إختر أفضل إجابة.

١. نعم

٢. لا

٣. تخطي هذا السؤال

**نهاية المسح**

#٣٢. شكراً جزيلا على وقتك! إذا كنت ترغب في معرفة المزيد عن فيروس كورونا، يرجى التحدث إلى أحد مقدمي الرّعاية الصّحية أو زيارة موارد موثوقة مثل
https://www.unicef.org/lebanon/ar/Covid19
