## Supplementary material for "Perceptions of and obstacles to SARS-CoV-2 vaccination among adults in Lebanon: a cross-sectional online survey": Survey English

**Consent and instructions:**
Thank you for your interest in “Shou Raa’yak?”, a survey about coronavirus and the coronavirus vaccine.

This research study will help researchers from the Modern University for Business and Science in Beirut and the University of California, Berkeley and Stanford University in the United States assist Lebanon’s response to coronavirus.

This survey is anonymous and does not collect any information that can determine your identity. Because this is a research study, we will inform you of the potential benefits and risks of participating, which are described in question #1. You will not be compensated for your participation.

If you are having symptoms or other health concerns, you should seek advice from a health care provider. If you are having an emergency, please stop this survey and find a doctor or hospital, or call 01/594459, the Ministry of Public Health Coronavirus Hotline.

#1. Please read the detailed informed consent document, available below. Do you affirm that you have read the informed consent document, understand the risks and benefits, and wish to participate in the research study?
[Image of the informed consent document will be embedded here]
1. Yes, I understand and wish to participate in the research study
2. No, I do not wish to participate in the research study

Branch point:
If “Yes,” proceed to question #2.
If “No,” proceed to question #32. (Thank you and further resources).

**Essential demographics:**

#2. How old are you (in years)? Please select from this dropdown list.

Branch point:
If number between 18-120, proceed to question #3.
If not number between 18-120, proceed to question #32 (Thank you and further resources).

#3. Do you currently live in Lebanon?
1. Yes
2. No

Branch point:
If “Yes,” proceed to question #4.
If “No,” proceed to question #32 (Thank you and further resources).

**Coronavirus Vaccine**

#4. The next questions are about health and coronavirus (the disease also referred to as "COVID-19," caused by the new virus "SARS-CoV-2"). Have you heard about coronavirus?
1. Yes
2. No

Branch point:
If "Yes", proceed to question #5.
If "No," proceed to question #32 (Thank you and further resources).

#5. Scientists have developed vaccines to protect individuals from coronavirus. If a vaccine becomes available to you, do you plan on receiving the coronavirus vaccine?
Choose the best single answer.
1. Yes
2. No
3. Not sure
4. Skip this question

Branch point:
If "Yes", proceed to question #6.
If "No", proceed to question #7.
If "Not sure", proceed to question #8.
If "Skip this question", proceed to question #11.

#6. Why do you plan to get the coronavirus vaccine if and when it becomes available to you?
Please type your response in a few sentences. If you do not wish to answer, simply leave the answer blank and advance to the next question.
[Free response]

Branch point:
Regardless of response, proceed to question #11.

#7. Why do you plan on **not** getting the coronavirus vaccine if and when it becomes available to you?
Please type your response in a few sentences. If you do not wish to answer, simply leave the answer blank and advance to the next question.
[Free response]

Branch point:
Regardless of response, proceed to question #9.

#8. Why are you **unsure** about getting the coronavirus vaccine if and when it becomes available?
Please type your response in a few sentences. If you do not wish to answer, simply leave the answer blank and advance to the next question.
[Free response]

Branch point:
Regardless of response, proceed to question #9.

#9. To be clear, there is no plan to pay people to take the coronavirus vaccine. However, imagine that if you were offered money, would that change your mind so that you decide to get the coronavirus vaccine?
1. Yes
2. No
3. Skip this question

Branch point:
If “Yes”, proceed to question #10.
If “No” or “Skip this question,” proceed to question #11.

#10. How much money would you need to receive so that you change your mind to get the coronavirus vaccine? Please select **all** the monetary amounts that would be enough for you to change your mind so that you get the coronavirus vaccine.
For example, imagine that you would take the vaccine if you received 25,000 LL, but would not take the vaccine if you were offered less than that. In this case, you would select answers 1, 2, 3, and 4.
Again, to be clear, there is no plan to pay people to take the coronavirus vaccine. This is a hypothetical question.
If you wish not to answer, please select “skip this question.”
[Select all that apply]
1. More than 100,000 LL
2. Between 76,000 LL and 100,000 LL
3. Between 51,000 LL and 75,000 LL
4. Between 21,000 LL and 50,000 LL
5. Between 11,000 LL and 20,000 LL
6. Between 1,000 LL and 10,000 LL
7. I will **not** get the coronavirus vaccine no matter how much money I am offered
8. Skip this question

#11. To be clear, it has not been determined whether the coronavirus vaccine will be free of cost to individuals getting vaccinated or if it will cost money. However, please imagine that you could purchase a vaccine. What is **the most** that you would be willing to pay of your own money (not including insurance) for a coronavirus vaccine when it becomes available?
Choose the best single answer.
1. Zero LL
2. Between 1,000 LL and 10,000 LL
3. Between 11,000 LL and 20,000 LL
4. Between 21,000 LL and 50,000 LL
5. Between 51,000 LL and 75,000 LL
6. Between 76,000 LL and 100,000 LL
7. More than 100,000 LL
8. I will **not** get the coronavirus vaccine even if it is free
9. Skip this question

#12. Where would you prefer to get a coronavirus vaccine?
Choose your top 3 preferred locations.

1. My doctor's office
2. Any doctor's office
3. A primary health center
4. A pharmacy
5. A hospital
6. A building close to my home that serves as a dedicated temporary vaccination clinic
7. A mobile van close to my home that serves as a dedicated temporary vaccination clinic
8. In my home, administered by a visiting certified vaccination provider
9. Other
10. I will **not** get the coronavirus vaccine regardless of available locations
11. Skip this question

    Branch point:
    If "9", proceed to question #13.
    If not "9", proceed to question #14.

#13. Please describe where you would prefer to get a coronavirus vaccine.
Please type your response in a few words. If you do not wish to answer, simply leave the answer blank and advance to the next question.
[Free response]

#14. If you have children under 18 years old, do you plan on having your children receive the coronavirus vaccine if and when it becomes available and is recommended by doctors for children?
Choose the best single answer.

1. Yes
2. No
3. Not sure
4. I do not have children under 18 years old
5. Skip this question

    Branch point:
    If "1", proceed to question #15.
    If "2", proceed to question #16.
    If "3", proceed to question #17.
    If "4" or "5", proceed to question #18.

#15. Why do you plan on having your children receive the coronavirus vaccine if and when it becomes available and recommended for children?
Please type your response in a few sentences. If you do not wish to answer, simply leave the answer blank and advance to the next question.
[Free response]

    Branch point:
    Regardless of response, proceed to question #18.

#16. Why do you plan on **not** having your children receive the coronavirus vaccine if and when it becomes available and recommended for children?
Please type your response in a few sentences. If you do not wish to answer, simply leave the answer blank and advance to the next question.

[Free response]

Branch point:
Regardless of response, proceed to question #18.

#17. Why are **you unsure** about having your children receive the coronavirus vaccine if and when it becomes available and recommended for children?
Please type your response in a few sentences. If you do not wish to answer, simply leave the answer blank and advance to the next question.
[Free response]

Branch point:
Regardless of response, proceed to question #18.

**Experience with coronavirus**

#18. In your understanding, how does someone become infected with coronavirus?
Choose the best single answer.
1. Eating raw food or untreated water
2. Being physically close to an infected person
3. Being bitten by an insect
4. Skip this question

#19. Have you ever been diagnosed with coronavirus, or thought you had coronavirus because of symptoms but were never officially diagnosed?
1. Yes
2. No
3. Skip this question

#20. Do you know someone in your family or a close friend who has had coronavirus?
1. Yes
2. No
3. Skip this question

#21. How frequently or infrequently do you wear a face mask when you are not at your home?
Choose the best single answer.
1. Never
2. Rarely
3. Sometimes
4. Most of the time
5. Always
6. Skip this question

#22. Where do you receive most of your information and news about coronavirus?
Choose your top 3 most **common** news sources.
1. Newspaper or magazine

2. Radio
3. Television
4. Social media (like Facebook, Twitter, YouTube, WhatsApp)
5. Internet, but not social media (like websites)
6. Talking to friends or family
7. Religious leaders
8. Skip this question

#23. Which news source do you trust the most for information and news about coronavirus?
Choose your top 3 most **trusted** news sources.
1. Newspaper or magazine
2. Radio
3. Television
4. Social media (like Facebook, Twitter, YouTube, WhatsApp)
5. Internet, but not social media (like websites)
6. Talking to friends or family
7. Religious leaders
8. Skip this question

**Additional demographics:**

#24. Which best describes you?
Choose the best single answer.
1. Female
2. Male
3. Other
4. Skip this question

#25. In which governorate do you live?
Choose the best single answer.
1. Baalbek-Hermel
2. Beqaa
3. Beirut
4. Mount Lebanon
5. South
6. Akkar
7. North
8. Nabatieh
9. Skip this question

#26. What is the highest level of education you have completed?
Choose the best single answer.
1. Never completed any formal school
2. Elementary school

3. High school
4. College or university
5. Graduate or professional school
6. Skip this question

#27. What is your religion?
Choose the best single answer.
1. Skip this question
2. Sunni
3. Shi'a
4. Druze
5. Other Muslim
6. Maronite
7. Catholic
8. Orthodox
9. Other Christian
10. Other religion or multiple religions
11. No religion

#28. What is your employment status?
Choose the best single answer.
1. Employed and exclusively working from home
2. Employed and physically going to work at least some of the time
3. Unemployed and looking for a job
4. Unemployed and not looking for a job
5. Retired
6. Homemaker
7. Student
8. Skip this question

#29. What is your estimated total household income in the year 2019?
Choose the best single answer.
1. Skip this question
2. Less than 1,000,000 LL
3. Between 1,000,000 LL and 1,999,999 LL
4. Between 2,000,000 LL and 9,999,999 LL
5. Between 10,000,000 LL and 19,999,999 LL
6. Between 20,000,000 LL and 39,999,999 LL
7. Between 40,000,000 LL and 69,999,999 LL
8. Between 70,000,000 LL and 99,999,999 LL
9. 100,000,000 LL or more

#30. What is your citizenship? If you are a citizen of multiple countries, you can choose more than one answer.

1. Lebanon
2. Syria
3. Palestine
4. Other Middle Eastern country (like Iraq, Egypt, Jordan, etc.)
5. Sub-Saharan African country (like Ethiopia, Kenya, etc.)
6. South Asian country (like Bangladesh, Sri Lanka, India, etc.)
7. Pacific Islands country (like Philippines, Indonesia, etc)
8. European or North American country (like France, Canada, etc.)
9. Other country
10. Skip this question

#31. Do you consider yourself a refugee living in Lebanon?
Choose the best single answer.
1. Yes
2. No
3. Skip this question

**Thank you/Link to resources at end of survey:**

#32. Thank you so much for your time! If you wish to learn more about coronavirus, please speak to a healthcare provider or visit trusted resources like this:
https://www.unicef.org/lebanon/ar/Covid19
