## Supplementary material for "Perceptions of and obstacles to SARS-CoV-2 vaccination among adults in Lebanon: a cross-sectional online survey": Tables and Figures included in main manuscript

### TABLES AND FIGURES FOR MAIN MANUSCRIPT

**Figure 1: Survey Recruitment and Outcomes**

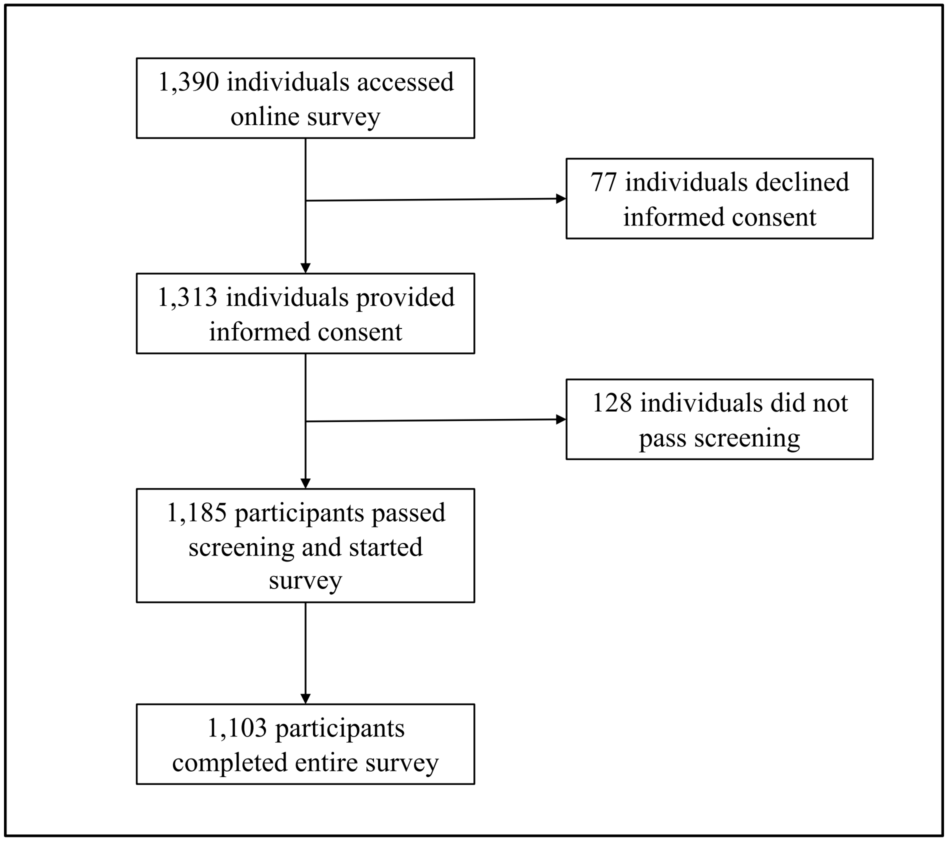

**Table 1: Participant Characteristics**

This includes all who passed screening. The first column includes all participants aggregated. The second column describes those who completed the survey in the first round, prior to initiation of vaccination in Lebanon on 13 February 2021. The third column describes those who completed the survey in the second round, after initiation of vaccination in Lebanon. Because participants were not forced to answer all questions, this results in a different denominator for each question.

LL: Lebanese Lira

NA: Not available in data source

[a]Participants could select multiple answers. Proportions were calculated using a denominator of all participants who selected an answer for the question.

[b]Unless otherwise noted, estimates were obtained from government source that excluded refugees and used 4.84 million (2018) as total population.

[c]Estimates were obtained from source that included refugees and used 6.86 million (2020) as total population.

| Characteristic | All Participants, n (%) | Round 1 Participants, n (%) | Round 2 Participants, n (%) | Estimates for Lebanese Population[b][1–5] |
|---|---|---|---|---|
| **Total Participants** | 1185 (100) | 840 (70.9) | 345 (29.1) | |
| **Gender** | | | | |
| Female | 685 (62.1) | 479 (60.1) | 206 (65.6) | 51.6% |
| Male | 388 (35.2) | 285 (36.1) | 103 (32.8) | 48.4% |
| Other | 7 (0.1) | 6 (0.8) | 1 (0.3) | NA |
| Skip this question | 23 (2.1) | 19 (2.4) | 4 (1.3) | |
| **Age** | | | | |
| 18-24 years old | 369 (31.1) | 246 (29.3) | 123 (35.7) | 15-24 years old: 17.5% |
| 25-34 years old | 376 (31.7) | 247 (29.4) | 129 (37.4) | 14.5% |
| 35-44 years old | 192 (16.2) | 144 (17.1) | 48 (13.9) | 11.8% |
| 45-54 years old | 126 (10.6) | 104 (12.4) | 22 (6.4) | 11.7% |
| 55-64 years old | 88 (7.4) | 69 (8.2) | 19 (5.5) | 9.6% |
| 65 years old or older | 34 (2.9) | 30 (3.6) | 4 (1.2) | 10.2% |

| | | | | |
|---|---|---|---|---|
| **Governorate** | | | | |
| Baalbek-Hermel | 21 (1.9) | 6 (0.8) | 15 (4.8) | 5.1% |
| Beqaa | 100 (9.1) | 47 (6.0) | 53 (16.9) | 6.2% |
| Beirut | 111 (10.1) | 89 (11.3) | 22 (7.0) | 7.1% |
| Mount Lebanon | 633 (57.4) | 559 (70.8) | 74 (23.6) | 42.3% |
| South | 72 (6.5) | 26 (3.3) | 46 (14.6) | 12.2% |
| Akkar | 13 (1.2) | 3 (0.4) | 10 (3.2) | 6.8% |
| North | 58 (5.3) | 21 (2.7) | 37 (11.8) | 13.3% |
| Nabatieh | 78 (7.1) | 22 (2.8) | 56 (17.8) | 7.9% |
| Skip this question | 17 (1.5) | 16 (2.0) | 1 (0.3) | |
| **Religion** | | | | |
| Christian | 242 (21.9) | 199 (25.2) | 43 (13.7) | 32.4% |
| Druze | 355 (32.2) | 308 (39.0) | 47 (15.0) | 4.5% |
| Shi'a | 136 (12.3) | 46 (5.8) | 90 (28.7) | 31.0% |
| Sunni | 124 (11.2) | 64 (8.1) | 60 (19.1) | 31.9% |
| No religion | 65 (5.9) | 48 (6.1) | 17 (5.4) | NA |
| Other | 6 (0.5) | 3 (0.4) | 3 (1.0) | 0.3% |
| Skip this question | 175 (15.9) | 121 (15.3) | 54 (17.2) | |
| **Highest Education Level** | | | | |
| Completed high school, technical school, or less | 200 (18.1) | 143 (18.1) | 57 (18.2) | 78.6% |
| Completed some college or more | 891 (80.9) | 637 (80.8) | 254 (80.9) | 21.4% |
| Skip this question | 11 (1.0) | 8 (1.0) | 3 (1.0) | |

| | | | | |
|---|---|---|---|---|
| **Employment** | | | | |
| Employed | 586 (51.5) | 412 (52.2) | 156 (49.7) | NA |
| Student | 179 (16.2) | 117 (14.8) | 62 (19.7) | NA |
| Unemployed, not seeking work | 145 (13.1) | 122 (15.5) | 23 (7.3) | NA |
| Unemployed, seeking work | 154 (14.0) | 101 (12.8) | 53 (16.9) | 33.0%[c] |
| Skip this question | 57 (5.2) | 37 (4.7) | 20 (6.3) | |
| **Annual Income (2019)** | | | | |
| < 1,000,000 LL | 122 (11.1) | 78 (9.9) | 44 (14.0) | Reliable data unavailable |
| 1,000,000 LL - 9,999,999 LL | 289 (26.2) | 199 (25.2) | 90 (28.7) | |
| 10,000,000 LL - 19,999,999 LL | 99 (9.0) | 67 (8.5) | 32 (10.2) | |
| 20,000,000 LL - 69,999,999 LL | 132 (12.0) | 100 (12.7) | 32 (10.2) | |
| >70,000,000 LL | 61 (5.5) | 49 (6.2) | 12 (3.8) | |
| Skip this question | 400 (36.3) | 296 (37.5) | 104 (33.1) | |
| **Citizenship[a]** | | | | |
| Lebanon | 1038 (94.1) | 757 (95.9) | 281 (89.5) | Lebanon: 79.8% |
| Syria | 28 (2.5) | 6 (0.8) | 6 (1.9) | Not citizen of Lebanon: 20.2% |
| Palestine | 17 (1.5) | 12 (1.5) | 12 (3.8) | |
| European or North American country | 40 (3.6) | 32 (4.1) | 32 (10.2) | |
| Other country | 23 (2.1) | 16 (2.0) | 16 (5.1) | |
| Skip this question | 12 (1.1) | 8 (1.0) | 8 (2.5) | |
| Multiple countries | 53 (4.8) | 40 (5.1) | 40 (12.7) | |
| **Refugee** | | | | |

| | | | | |
|---|---|---|---|---|
| Yes | 56 (5.1) | 29 (3.7) | 27 (8.6) | 21.9%[c] |
| No | 1015 (92.0) | 737 (93.4) | 278 (88.5) | 78.1%[c] |
| Skip this question | 32 (2.9) | 23 (2.9) | 9 (2.9) | |

**Table 2: Intentions about vaccination by sociodemographic characteristics**

This table shows intention to vaccinate by demographic characteristic for everyone who answered the intention to vaccinate question. For the analysis of each characteristic, we omitted participants who skipped the question, unless >10% of participants for that question skipped the question, in which case those who skipped the characteristic question were included in the analysis. We then calculated the proportion of each characteristic subcategory by intention to vaccinate, calculating Wilson Score confidence intervals.

[a]The survey allowed participants to choose multiple answers for this characteristic; consequently, the sum of all subcategories does not equal the number of all participants who answered the question.

| Characteristic | Intend to receive vaccine when available | % All Participants [95% CI] |
|---|---|---|
| **All participants** | | |
| | Yes | 46.1% [43.2%-49.0%] |
| | No | 19.0% [16.8%-21.4%] |
| | Unsure | 34.0% [31.3%-36.8%] |
| **Gender** | | |
| Female | | |
| | Yes | 42.8% [39.1%-46.6%] |
| | No | 21.1% [18.1%-24.4%] |
| | Unsure | 36.1% [32.5%-39.8%] |
| Male | | |
| | Yes | 55.6% [50.5%-60.6%] |
| | No | 15.6% [12.2%-19.7%] |
| | Unsure | 28.8% [24.4%-33.7%] |
| Other | | |
| | Yes | 33.3% [6.0%-75.9%] |
| | No | 50.0% [18.8%-81.2%] |
| | Unsure | 16.7% [0.9%-63.5%] |

**Age**

| | | |
|---|---|---|
| 18-24 years old | | |
| | Yes | 39.6% [34.5%-44.8%] |
| | No | 22.5% [18.4%-27.2%] |
| | Unsure | 37.9% [32.9%-43.1%] |
| 25-34 years old | | |
| | Yes | 45.4% [40.3%-50.6%] |
| | No | 23.1% [19.0%-27.8%] |
| | Unsure | 31.5% [26.8%-36.5%] |
| 35-44 years old | | |
| | Yes | 50.8% [43.5%-58.0%] |
| | No | 14.1% [9.7%-20.0%] |
| | Unsure | 35.1% [28.4%-42.3%] |
| 45-54 years old | | |
| | Yes | 51.2% [42.1%-60.2%] |
| | No | 12.0% [7.1%-19.3%] |
| | Unsure | 36.8% [28.5%-45.9%] |
| 55-64 years old | | |
| | Yes | 60.2% [49.2%-70.3%] |
| | No | 12.5% [6.7%-21.7%] |
| | Unsure | 27.3% [18.6%-38.0%] |
| 65 years old or older | | |
| | Yes | 55.9% [38.1%-72.4%] |
| | No | 11.8% [3.8%-28.4%] |
| | Unsure | 32.4% [18.0%-50.6%] |

**Governorate**

| | | |
|---|---|---|
| Baalbek-Hermel | | |
| | Yes | 50.0% [29.9%-70.1%] |
| | No | 15.0% [4.0%-38.9%] |
| | Unsure | 35.0% [16.3%-59.1%] |
| Beqaa | | |
| | Yes | 59.2% [48.8%-68.9%] |
| | No | 12.2% [6.8%-20.8%] |
| | Unsure | 28.6% [20.1%-38.7%] |
| Beirut | | |
| | Yes | 53.2% [43.5%-62.6%] |
| | No | 16.2% [10.1%-24.7%] |
| | Unsure | 30.6% [22.4%-40.2%] |
| Mount Lebanon | | |
| | Yes | 42.2% [38.4%-46.2%] |
| | No | 22.8% [19.6%-26.3%] |
| | Unsure | 35.0% [31.3%-38.8%] |
| South | | |
| | Yes | 57.1% [44.8%-68.7%] |
| | No | 10.0% [4.5%-20.1%] |
| | Unsure | 32.9% [22.4%-45.2%] |
| Akkar | | |
| | Yes | 84.6% [53.7%-97.3%] |
| | No | 7.7% [0.4%-37.9%] |
| | Unsure | 7.7% [0.4%-37.9%] |
| North | | |
| | Yes | 46.6% [33.5%-60.0%] |
| | No | 17.2% [9.0%-29.9%] |
| | Unsure | 36.2% [24.3%-49.9%] |
| Nabatieh | | |

| | | |
|---|---|---|
| | Yes | 50.6% [39.1%-62.1%] |
| | No | 18.2% [10.6%-29.0%] |
| | Unsure | 31.2% [21.4%-42.9%] |
| **Religion** | | |
| Christian | | |
| | Yes | 60.3% [53.8%-66.4%] |
| | No | 9.5% [6.2%-14.1%] |
| | Unsure | 30.2% [24.5%-36.4%] |
| Druze | | |
| | Yes | 35.6% [30.6%-40.9%] |
| | No | 25.4% [21.0%-30.3%] |
| | Unsure | 39.0% [33.9%-44.3%] |
| Shi'a | | |
| | Yes | 48.5% [39.8%-57.3%] |
| | No | 16.4% [10.8%-24.0%] |
| | Unsure | 35.1% [27.2%-43.8%] |
| Sunni | | |
| | Yes | 57.4% [48.1%-66.2%] |
| | No | 21.3% [14.6%-29.8%] |
| | Unsure | 21.3% [14.6%-29.8%] |
| No religion | | |
| | Yes | 58.5% [45.6%-70.3%] |
| | No | 16.9% [9.1%-28.7%] |
| | Unsure | 24.6% [15.1%-37.1%] |
| Other | | |
| | Yes | 20.0% [1.1%-70.1%] |
| | No | 0.0% [0.0%-53.7%] |
| | Unsure | 80.0% [29.9%-99.0%] |

| | | |
|---|---|---|
| Skip this question | | |
| | Yes | 39.9% [32.6%-47.6%] |
| | No | 24.9% [18.8%-32.1%] |
| | Unsure | 35.3% [28.3%-42.9%] |
| **Education** | | |
| Completed high school, technical school, or less | | |
| | Yes | 42.1% [35.2%-49.4%] |
| | No | 21.3% [16.0%-27.8%] |
| | Unsure | 36.5% [30.0%-43.7%] |
| Completed some college or more | | |
| | Yes | 48.8% [45.4%-52.1%] |
| | No | 18.5% [16.0%-21.3%] |
| | Unsure | 32.7% [29.7%-35.9%] |
| **Employment** | | |
| Employed | | |
| | Yes | 51.6% [47.4%-55.8%] |
| | No | 17.1% [14.2%-20.6%] |
| | Unsure | 31.3% [27.5%-35.3%] |
| Student | | |
| | Yes | 43.0% [35.7%-50.6%] |
| | No | 21.2% [15.6%-28.1%] |
| | Unsure | 35.8% [28.8%-43.3%] |

| | |
|---|---|
| Unemployed, not seeking work | |
| Yes | 42.1% [34.0%-50.6%] |
| No | 18.6% [12.8%-26.1%] |
| Unsure | 39.3% [31.4%-47.8%] |
| Unemployed, seeking work | |
| Yes | 41.4% [33.6%-49.7%] |
| No | 23.0% [16.8%-30.7%] |
| Unsure | 35.5% [28.1%-30.7%] |
| **Annual Income** | |
| < 1,000,000 LL | |
| Yes | 34.7% [26.4%-44.0%] |
| No | 25.6% [18.3%-34.5%] |
| Unsure | 39.7% [31.0%-49.0%] |
| 1,000,000 LL - 9,999,999 LL | |
| Yes | 45.6% [39.8%-51.6%] |
| No | 18.8% [14.6%-23.9%] |
| Unsure | 35.5% [30.1%-41.4%] |
| 10,000,000 LL - 19,999,999 LL | |
| Yes | 47.5% [37.4%-57.7%] |
| No | 24.2% [16.4%-34.1%] |
| Unsure | 28.3% [19.9%-38.4%] |
| 20,000,000 LL - 69,999,999 LL | |
| Yes | 63.4% [54.4%-71.5%] |
| No | 9.2% [5.0%-15.8%] |
| Unsure | 27.5% [20.2%-36.1%] |
| >70,000,000 LL | |
| Yes | 68.8% [55.6%-79.8%] |
| No | 6.6% [2.1%-16.7%] |

| | | |
|---|---|---|
| | Unsure | 24.6% [14.8%-37.6%] |
| Skip this question | | |
| | Yes | 42.9% [38.0%-48.0%] |
| | No | 22.7% [18.8%-27.2%] |
| | Unsure | 34.3% [29.7%-39.3%] |
| **Citizenship**[a] | | |
| Lebanon | | |
| | Yes | 47.2% [44.2%-50.3%] |
| | No | 19.1% [16.7%-21.6%] |
| | Unsure | 33.7% [30.8%-36.7%] |
| Syria | | |
| | Yes | 40.7% [23.0%-61.0%] |
| | No | 37.0% [20.1%-57.5%] |
| | Unsure | 22.2% [9.4%-42.7%] |
| Palestine | | |
| | Yes | 58.8% [33.5%-80.6%] |
| | No | 11.8% [2.1%-37.7%] |
| | Unsure | 29.4% [11.4%-56.0%] |
| European or North American country | | |
| | Yes | 62.5% [45.8%-76.8%] |
| | No | 12.5% [4.7%-27.6%] |
| | Unsure | 25.0% [13.2%-41.5%] |
| Other country | | |
| | Yes | 31.8% [14.7%-54.8%] |
| | No | 18.2% [6.0%-41.0%] |
| | Unsure | 50.0% [30.7%-69.3%] |

| | | |
|---|---|---|
| Multiple countries | | |
| | Yes | 54.7% [40.6%-68.2%] |
| | No | 13.2% [5.9%-26.0%] |
| | Unsure | 32.1% [20.3%-46.4%] |
| **Refugee** | | |
| Yes | | |
| | Yes | 50.0% [37.1%-62.9%] |
| | No | 25.9% [15.4%-39.9%] |
| | Unsure | 24.1% [13.9%-37.9%] |
| No | | |
| | Yes | 46.9% [43.8%-50.0%] |
| | No | 19.0% [16.7%-21.6%] |
| | Unsure | 34.1% [31.2%-37.1%] |
| **Completed Survey After Initiation of Vaccination in Lebanon** | | |
| Yes | | |
| | Yes | 56.9% [51.5%-62.2%] |
| | No | 12.7% [9.4%-16.8%] |
| | Unsure | 30.4% [25.6%-35.6%] |
| No | | |
| | Yes | 42.3% [38.9%-45.7%] |
| | No | 21.8% [19.1%-24.8%] |
| | Unsure | 35.9% [32.7%-39.3%] |

**Figure 2: Intention to receive SARS-CoV-2 vaccine by Age Group and Gender**
In these graphs, the sample population is stratified by age group and gender. For each stratum, the proportion (as a percentage) that indicated that they intend to receive the vaccine, do not intend to receive the vaccine, and are unsure about receiving the vaccine is displayed in the corresponding cell in the top, middle, and bottom graphs, respectively. This analysis only includes participants who provided age, gender, and their intentions about vaccination. As only 7 participants identified as "other" gender, only participants identifying as "male" or "female" were included.

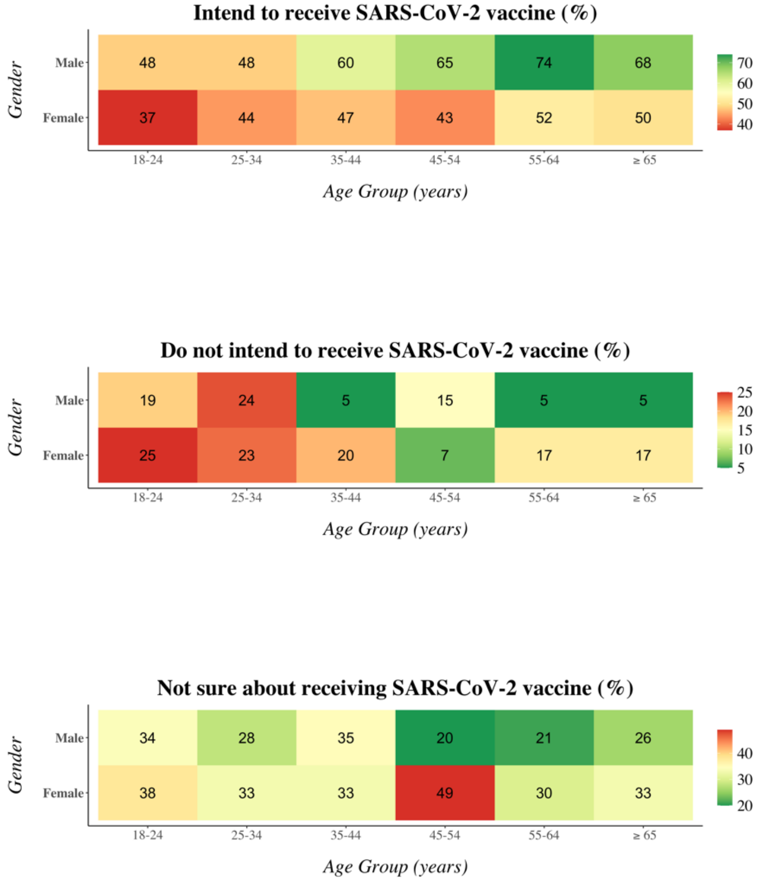

**Table 3: Intentions about vaccination by experiences with COVID-19**

This shows intentions to vaccinate broken down by knowledge of and experience with COVID-19 for everyone who answered the intention to vaccinate question. For the analysis of each characteristic, we omitted participants who skipped the question, unless >10% of participants for that question skipped the question, in which case those who skipped the characteristic question were included in the analysis. We then calculated the proportion of each characteristic subcategory by intention to vaccinate, calculating Wilson Score confidence intervals.

[a]"Correct Knowledge" indicated a correct response to a multiple choice question asking, "In your understanding, how does someone become infected with coronavirus? Choose the best single answer." The correct response was "Being physically close to an infected person." Incorrect responses were "Eating raw food or untreated water," and "Being bitten by an insect."

[b]The survey allowed participants to choose multiple answers for this characteristic; consequently, the sum of all subcategories does not equal the number of all participants who answered the question.

| Characteristic | Intend to receive vaccine when available | All Participants [95% CI] |
|---|---|---|
| **Correct Knowledge of Transmission[a]** | | |
| Yes | | n = 1076 (99.1%) |
| | Yes | 48.0% [45.1%-51.1%] |
| | No | 18.1% [15.9%-20.6%] |
| | Unsure | 33.8% [31.0%-36.8%] |
| No | | n = 10 (0.9%) |
| | Yes | 50.0% [23.7%-76.3%] |
| | No | 40.0% [13.7%-72.6%] |
| | Unsure | 10.0% [0.5%-45.9%] |
| **Personal history of COVID-19** | | |
| Yes | | n = 321 (28.9%) |
| | Yes | 42.9% [37.5%-48.6%] |

| | | |
|---|---|---|
| | No | 21.6% [17.3%-26.6%] |
| | Unsure | 35.4% [30.2%-41.0%] |
| No | | n = 790 (71.1%) |
| | Yes | 48.8% [45.2%-52.3%] |
| | No | 18.5% [15.9%-21.5%] |
| | Unsure | 32.7% [29.4%-36.1%] |
| **Close acquaintance with history of COVID-19** | | |
| Yes | | n = 1056 (94.2%) |
| | Yes | 48.0% [44.9%-51.0%] |
| | No | 18.8% [16.5%-21.3%] |
| | Unsure | 33.3% [30.4%-36.2%] |
| No | | n = 65 (5.8%) |
| | Yes | 33.3% [22.2%-46.4%] |
| | No | 30.2% [19.6%-43.2%] |
| | Unsure | 36.5% [25.0%-49.6%] |
| **Mask Wearing Outside Home** | | |
| Always or Most of the Time | | n = 1011 (91.2%) |
| | Yes | 50.0% [46.8%-53.1%] |
| | No | 16.3% [14.1%-18.8%] |
| | Unsure | 33.7% [30.8%-36.8%] |
| Sometimes, Rarely, or Never | | n = 97 (8.8%) |
| | Yes | 18.6% [11.7%-28.0%] |
| | No | 48.5% [38.3%-58.8%] |

| | | |
|---|---|---|
| | Unsure | 33.0% [24.0%-43.4% |
| **Top 3 News Sources**[b] | | |
| Printed newspaper or magazine | | n = 86 (7.8%) |
| | Yes | 64.0% [52.8%-73.8%] |
| | No | 9.3% [4.4%-18.0%] |
| | Unsure | 26.7% [18.0%-37.6%] |
| Radio | | n = 33 (3.0%) |
| | Yes | 69.7% [51.1%-83.8%] |
| | No | 15.2% [5.7%-32.7%] |
| | Unsure | 15.2% [5.7%-32.7%] |
| Television | | n = 718 (65.1%) |
| | Yes | 47.2% [43.5%-50.9%] |
| | No | 17.1% [14.5%-20.1%] |
| | Unsure | 35.7% [32.2%-39.3%] |
| Social media (like Facebook, Twitter, YouTube, WhatsApp) | | n = 670 (60.1%) |
| | Yes | 43.7% [39.9%-47.6%] |
| | No | 19.4% [16.5%-22.6%] |
| | Unsure | 36.9% [33.2%-40.7%] |
| Internet, but not social media (like websites) | | n = 609 (55.2%) |
| | Yes | 53.5% [49.5%-57.5%] |
| | No | 17.4% [14.5%-20.7%] |
| | Unsure | 29.1% [25.5%-32.9%] |

| | | |
|---|---|---|
| Talking to friends or family | | n = 282 (25.6%] |
| | Yes | 37.6% [32.0%-43.6%] |
| | No | 24.1% [19.3%-29.6%] |
| | Unsure | 38.3% [32.6%-44.3%] |
| Religious leaders | | n = 6 (0.5%) |
| | Yes | 16.7% [0.9%-63.5%] |
| | No | 83.3% [36.5%-99.1%] |
| | Unsure | 0.0% [0.0%-48.3%] |

**Figure 3: Preferred vaccination locations**
Percentages of each response were calculated by dividing the number of participants who selected a response by the total number of participants who answered this question.
*The survey allowed participants to choose multiple answers for this question; consequently, the sum of all subcategories does not equal the number of all participants who answered the question.
^If participants selected "Other," they could write in their own response. The three responses in parenthesis (Lebanese Red Cross, Military Primary Health Center, other health organization) encompass the written-in responses.

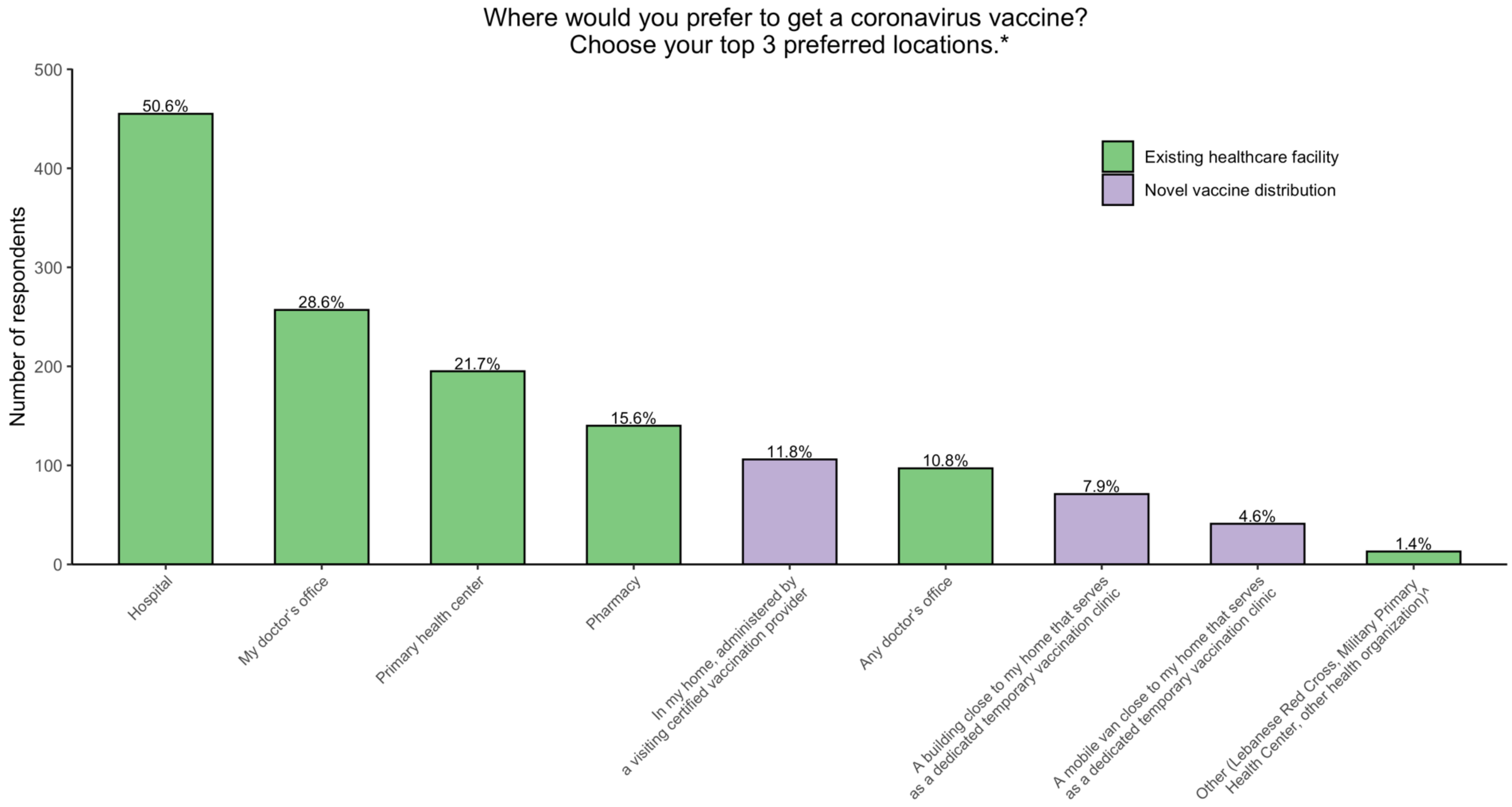

**Figure 4: Most common and most trusted news sources about coronavirus**
This figure summarizes responses to two separate questions, the first asking which news sources participants most commonly used, and the second asking which news sources they trusted most. Percentages of each response were calculated by dividing the number of participants who selected a response by the total number of participants who answered that question.
*The survey allowed participants to choose multiple answers for these questions; consequently, the sum of all subcategories does not equal the number of all participants who answered the questions.

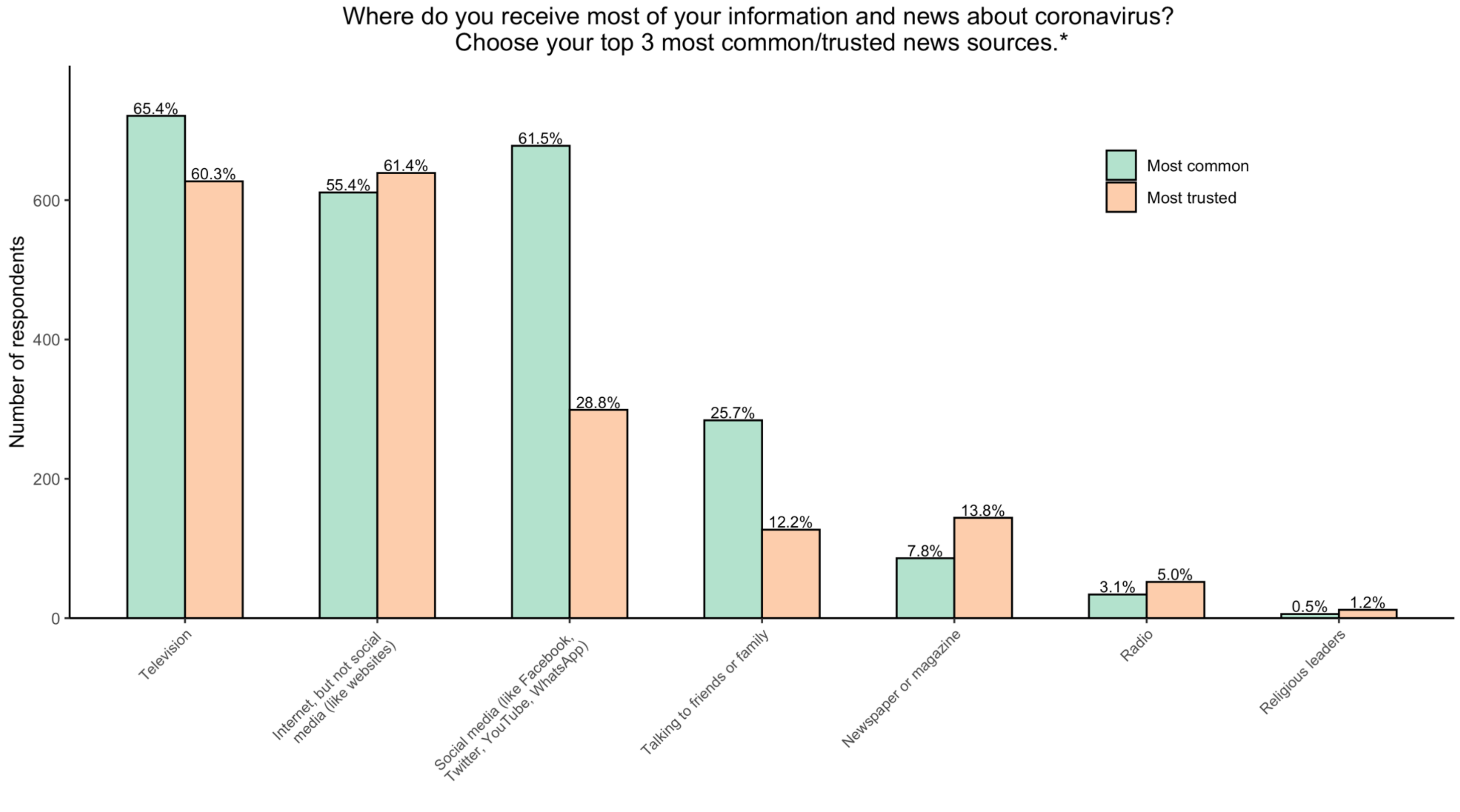

**Table 4: Summary of coded open-ended responses describing motivations for intentions about vaccination**
This shows the most frequently observed codes in the optional open-ended responses to questions asking about motivations behind deciding to vaccinate, deciding not to vaccinate, and being uncertain about vaccination. For each group (intending to vaccinate, not intending to vaccinate, and uncertainty about vaccination), the denominator was the total number of participants in the group who provided a free-response to the question.
[a]"Most frequent codes" were observed in ≥15% of responses to the question.
[b]"Less frequent codes" were observed in 3%-15% of responses to the question.

| TABLE 4: SUMMARY OF CODED OPEN-ENDED RESPONSES DESCRIBING MOTIVATIONS | | |
|---|---|---|
| | **Most frequent codes (descending frequency)** | **Less frequent codes (descending frequency)** |
| **Motivations for intending to vaccinate** | To protect myself<br>To protect and limit spread among the public<br>To end the pandemic and return to normalcy<br>To protect my family | I do not have another better choice or solution<br>I trust it based on science and research<br>I want to help achieve "herd immunity"<br>I have other medical conditions that make my risk of illness higher<br>My job puts me at risk of contracting COVID-19 |
| **Motivations for not intending to vaccinate** | I am concerned about safety and limited testing<br>I am concerned about potential side effects<br>Nonspecific mistrust of the SARS-CoV-2 vaccines<br>I am worried that the vaccines are not effective | I do not trust the Lebanese government<br>I do not trust Lebanese healthcare and/or distribution systems<br>COVID-19 and/or vaccines are a scam/conspiracy to make money<br>General vaccine hesitancy; I am against all vaccines<br>Problems or concerns with ingredients |

| | | |
|---|---|---|
| | | I do not need the vaccine because I already had COVID-19<br>COVID-19 is not a threat to me or in general<br>Risks of vaccination are not worth the potential benefits<br>Society and science in general do not know enough about COVID-19 (causes, symptoms, effects) |
| **Motivations for uncertainty about vaccination** | I am concerned about safety and limited testing<br>I am concerned about potential side effects<br>I am worried that the vaccines are not effective | I do not think I need the vaccine because I already had COVID-19<br>I do not trust the Lebanese government<br>Nonspecific mistrust of the SARS-CoV-2 vaccines<br>I do not have enough information about the vaccines<br>I want more time or more information before I decide which vaccine to take<br>I have received conflicting information about the vaccine from one or more sources<br>I have concerns about mRNA technology<br>I do not trust Lebanese healthcare and/or distribution systems<br>I think the vaccines in Lebanon will not be the true coronavirus vaccine, or that they will be tampered with |

### TABLES AND FIGURES FOR APPENDIX

**Table 5: Logistical considerations about vaccination**
This shows responses to questions about logistical considerations about vaccination. For the analysis of each characteristic, we omitted participants who skipped the question, unless >10% of participants for that question skipped the question, in which case those who skipped the characteristic question were included in analysis.
[a]The survey allowed participants to choose multiple answers for this question; consequently, the sum of all subcategories does not equal the number of all participants who answered the question.
[b]Analysis for this question excludes those who skipped the question or who selected, "I will not get the coronavirus vaccine regardless of available locations."
[c]If participants selected "Other," they could write in their own response. The three responses in parenthesis (Lebanese Red Cross, Military Primary Health Center, other health organization) encompass the written-in responses.

| Logistical Consideration | Participants, n (%) |
|---|---|
| **Among participants who did not intend to get vaccinated or were unsure about receiving a vaccine: Would a monetary incentive change your mind so that you take a vaccine?** | |
| Yes | 10 (1.7%) |
| No | 589 (98.3%) |

| | |
|---|---|
| **Among all participants: What is the most of your own money that you would be willing to pay for a coronavirus vaccine?** | |
| Zero LL | 201 (19.4%) |
| Between 1,000 LL and 10,000 LL | 86 (8.3%) |
| Between 11,000 LL and 20,000 LL | 120 (11.6%) |
| Between 21,000 LL and 50,000 LL | 147 (14.2%) |
| Between 51,000 LL and 75,000 LL | 64 (6.2%) |
| Between 76,000 LL and 100,000 LL | 119 (11.5%) |
| More than 100,000 LL | 113 (10.9%) |
| I will not get the coronavirus vaccine even if it is free | 186 (18.0%) |
| **Where would you prefer to get a coronavirus vaccine? Choose your top 3 preferred locations.[a,b]** | |
| My doctor’s office | 257 (28.6%) |
| Any doctor’s office | 97 (10.8%) |
| A primary health center | 195 (21.7%) |
| A pharmacy | 140 (15.6%) |
| A hospital | 455 (50.6%) |

| | |
|---|---|
| A building close to my home that serves as a dedicated temporary vaccination clinic | 71 (7.9%) |
| A mobile van close to my home that serves as a dedicated temporary vaccination clinic | 41 (4.6%) |
| In my home, administered by a visiting certified vaccination provider | 106 (11.8%) |
| Other (Lebanese Red Cross, Military Primary Health Center, other health organization)[c] | 13 (1.4%) |
| **Where do you receive most of your information and news about coronavirus? Choose your top 3 most common news sources.[a]** | |
| Newspaper or magazine | 86 (7.8%) |
| Radio | 34 (3.1%) |
| Television | 721 (65.4%) |
| Social media (like Facebook, Twitter, YouTube, WhatsApp) | 678 (61.5%) |
| Internet, but not social media (like websites) | 611 (55.4%) |
| Talking to friends or family | 284 (25.7%) |
| Religious leaders | 6 (0.5%) |

| **Which news source do you trust the most for information and news about coronavirus? Choose your top 3 most trusted news sources.**[a] | |
|---|---|
| Newspaper or magazine | 144 (13.8%) |
| Radio | 52 (5.0%) |
| Television | 627 (60.3%) |
| Social media (like Facebook, Twitter, YouTube, WhatsApp) | 299 (28.8%) |
| Internet, but not social media (like websites) | 639 (61.4%) |
| Talking to friends or family | 127 (12.2%) |
| Religious leaders | 12 (1.2%) |

[1] Lebanon's Central Administration for Statistics (CAS), International Labour Organization. Labour Force and Household Living Conditions Survey (LFHLCS) in Lebanon 2018–2019. 2019.

[2] Central Intelligence Agency USA. Lebanon - The World Factbook n.d. https://www.cia.gov/the-world-factbook/countries/lebanon/ (accessed May 4, 2021).

[3] United Nations World Food Programme. Assessing the Impact of the Economic and COVID-19 Crises in Lebanon - June 2020. 2020.
[4] World Health Organization. Regional Office for the Eastern Mediterranean. Country cooperation strategy: Lebanon: 2019-2023. 2018.
[5] The World Bank. Lebanon | Data. 2021 n.d. https://data.worldbank.org/country/lebanon (accessed May 4, 2021).
